# Future HIV epidemic trajectories in South Africa and long-term consequences of reductions in general HIV testing: a mathematical modelling study

**DOI:** 10.1101/2023.12.19.23300231

**Authors:** Stefan P Rautenbach, Lilith K Whittles, Gesine Meyer-Rath, Lise Jamieson, Thato Chidarikire, Leigh F Johnson, Jeffrey W Imai-Eaton

**Author notes:** These authors contributed equally (Leigh F Johnson, Jeffrey W Imai-Eaton).

## Abstract

**Background:** Following successful intensive interventions to rapidly increase HIV awareness, antiretroviral therapy (ART) coverage, and viral suppression, HIV programmes in eastern and southern Africa must now consider whether to scale-back certain programmes, such as widespread general population HIV testing services (general HTS), without risking a resurging epidemic or substantially increasing long-term ART need through slowed incidence declines.

**Methods:** We used a mathematical model (Thembisa) to project the South African HIV epidemic to 2100 under current epidemiologic and programmatic conditions. We assessed the epidemiological impact and cost of implementing general HTS reductions at different times between 2025 and 2050, while maintaining antenatal, symptom-based, and risk-based testing modalities and other HIV prevention. We considered how future uncertainty interacted with testing reductions by assuming positive or negative changes in ART interruption rates and condom usage over 2025–2035.

**Findings:** Under the status quo scenario, HIV incidence (15-49 years) steadily declined from 4.95/1000 (95% CI: 4.40–5.34) in 2025 to 0.14/1000 (0.05–0.31) in 2100, attaining <1/1000 in 2055 (2051–2060). When general HTS was scaled-back in 2025, incidence continued declining, but time to <1/1000 was delayed by 5, 13, and 35 years for a 25%, 50%, or 75% reduction in general HTS, and not attained by 2100 with full cessation. Reducing general HTS by 25% to 100% from 2025 resulted in 10% (8–12%) to 65% (53–77%) more new HIV infections and 7% (5–8%) to 46% (38–53%) more AIDS-related deaths over 50 years. Delaying general HTS reductions for 5 to 25 years mitigated some impacts. HIV testing accounted for only 5% of total programmatic costs at baseline. Reducing testing modestly reduced short-term total costs, but increased long-term costs. Changes in ART interruption rates and condom usage levels affected incidence decline rates and general HTS levels required to control transmission but did not cause rapid resurgent incidence.

**Interpretation:** Scaling-back general HTS did not result in resurging HIV infections, but it delayed attainment of incidence reduction targets and increased long-term expected infections, deaths, ART provision, and costs. HIV programmes face decisions balancing near-term health system resource savings by reducing intensive HIV programmes with epidemic control objectives over several decades.

**Funding:** BMGF, Wellcome, UKRI

## Introduction

Over the past decade, HIV programmes in eastern and southern Africa have dramatically expanded HIV testing, access to treatment, and other interventions to rapidly increase population viral suppression and reduce new HIV infections and AIDS-related deaths, aligned with UNAIDS 95-95-95 targets^1,2^ and commitments to end AIDS as a public health threat by 2030.^1,3,4^ These intensive programmes succeeded at increasing viral suppression in the region from 46% in 2015 to 73% in 2021, with five countries already attaining the 95-95-95 targets of 86% suppression among people living with HIV (PLWH).^2^

Now, many programmes are considering how to evolve to sustain high viral suppression and low and declining new infections for decades to come. This includes whether, when, and by how much to scale-back resource intensive vertical programmes without risking resurgence of HIV infections or deaths or stagnating incidence declines adding to long-term treatment costs. Several countries, including Lesotho, Uganda, and Kenya, have recently substantially reduced general population HIV testing services (general HTS) testing.^5^ Long-term programme management decisions should consider robustness to future uncertainty about incidence dynamics. Epidemiologic uncertainties may include observable programme outcomes to which strategies could respond dynamically, such as improvement or deterioration in antiretroviral therapy (ART) retention and viral suppression, or harder-to-monitor determinants like changes in population risk-behaviour, such as changes in sexual partnering or decreased condom use.

South Africa, with 7.8 million PLWH (almost 1 in 5 adults),^6^ performs over 14 million HIV tests each year, which will increase to 20 million by 2039 if current testing trends continue.^7^ New HIV infections in South Africa have declined since 1997, including by 58% since 2010.^6^ Initial declines were attributed largely to increased condom usage,^8,9^ and more recently primarily due to scaling-up ART coverage via extensive HIV testing and expanded eligibility, along with other prevention such as voluntary medical male circumcision (VMMC).^9,10^ General HTS (as opposed to risk-targeted, symptomatic, or antenatal testing) constitutes 80% of all HIV tests, and may be scaled back as there are fewer untreated PLWH in the population.^9^

We used a mathematical model of HIV transmission in South Africa^11^ to simulate long-term epidemic trajectories to 2100 under current trends in testing rates, ART linkage and interruption, and current levels of other prevention interventions. We assessed how managed reductions to general HTS by different levels and time intervals affected ability to maintain declining incidence, the expected additional HIV infections and AIDS-related deaths over 50-years, and impact on HIV programme costs. Finally, we assessed the risk of future resurgence due to uncertainty in future observable programmatic outcomes, through ART retention, or underlying changes in sexual risk behaviour, such as condom-use, under alternative testing strategies.

## Methods

### Mathematical model

Thembisa is a deterministic compartmental model of HIV transmission and demography in South Africa, used for national HIV estimates and policy development, including estimates reported by UNAIDS.^9,11^ The model is calibrated to national data on HIV prevalence, mortality, sexual partnership and risk behaviour, and HIV programme testing, treatment, and prevention provision using a Bayesian approach.^11^ We used the Thembisa 4.5 model updated in 2022.

Model details and calibration are described elsewhere.^11^ Briefly, the population is stratified by demographic groups (age and sex), behavioural risk (sexual experience, marital status, propensity for partner concurrency, engagement in commercial sex work), and HIV status, with the HIV-positive population stratified by CD4 count, HIV testing history, and ART usage. The HIV epidemic is initialised in 1985. HIV transmission depends on the HIV-positive partner’s CD4 count and infection stage, prevention method use (condoms, preexposure prophylaxis [PrEP]), and circumcision status for HIV-negative male partners.

HIV diagnosis and linkage to ART occurs via six routes: (i) testing of pregnant women at antenatal clinics, (ii) testing prompted by opportunistic infections, (iii) testing of HIV-exposed infants, (iv) testing of partners of PLWH through passive partner notification, (v) self-testing, and (vi) other or ‘general’ HTS. General HTS included HIV testing of asymptomatic, non-pregnant adults either provider-or self-initiated, or initiated by insurance requirements.

### Scenario analysis

We projected the South African HIV epidemic to 2100 assuming continuation of current demographic trends and previously inferred trends in condom-use, ART initiation and retention, and HIV testing rates (Table S1), using 1000 calibrated model parameter sets to incorporate parametric uncertainty. HIV self-testing rates were low before 2021 and assumed to be zero from 2021 onwards. Uptake rates of other prevention interventions, including condom use, VMMC, and PrEP among key and high-risk populations were assumed to continue at 2025 levels.

To assess the potential impact of future reductions in general HTS, we simulated the epidemic assuming reductions between 0-100% compared to the 2021 testing rate (Table S2). Testing reductions were implemented at 5-year time intervals between 2025 and 2050. Reduction in general HTS was implemented over 2 years and maintained at the lower rate until 2100. Antenatal, symptom-based, and passive partner notification testing were maintained at 2021 rates.

In addition to reducing general HTS, we assessed the impact of future epidemiologic uncertainty by varying the rate of treatment interruption and sexual risk environment over 10 years from 2025–2035. We varied the ART interruption rate in scenarios ranging from a 14% annual reduction to a 14% annual increase relative the 2021 baseline (Table S3). This corresponded to ART coverage in 2035 between 50% and 92% versus 77% under status quo. Similarly, we modelled future changes in sexual risk by varying the odds of condom-use from a 14% reduction to a 14% increase over 2025–2035 relative the 2021 baseline (Table S4). Modelled scenarios considered condom-use (percentage of total sex acts in which a condom is used) between 14% and 58% in 2035, compared to 34% under status quo.

Model outputs were annual HIV incidence rate per 1000, prevalence among adults 15-49 years, HIV testing volume, treatment coverage, new infections, and AIDS-deaths among adults 15+ years. We calculated the time to reach HIV incidence (15-49 years) below 1 per 1000 (<1/1000), a proposed threshold for ‘virtual HIV elimination’,^12^ and additional HIV infections and AIDS-deaths over a 50-year time horizon from when general HTS was reduced. We estimated annual and cumulative HIV testing and treatment programme costs of alternative scenarios using previously applied methods.^7,13^ Costs were estimated from the provider’s perspective (South African government) and presented in 2023 United States dollar (US$). Costs were included for HIV testing (general HTS: US$3.63 per HIV-negative test, $5.20 for HIV-positive), annual ART ($272 first-year, $171 per subsequent year, $300 per year for second-line treatment), and in-patient and palliative care costs (ranging $37 to $402 per patient year depending on ART and CD4 category; see Table S5). We calculated the difference in annual testing and treatment costs over time and cumulative costs after 5, 10, 25, and 50 years discounted at 0%, 3%, and 6% per annum.

## Results

When maintaining current programme levels and risk environment (‘status quo’ scenario), adult (15-49 years) HIV incidence in South Africa was projected to continue steadily declining from 4.95 per 1000 (95% CI: 4.40–5.34) in 2025 to 0.14 per 1000 (0.05–0.31) in 2100 (Figure 1A), reducing below 1/1000 in 2055 (2051–2060). HIV prevalence among adults (15-49 years) declined from 17.7% (16.8–18.2%) in 2025 to 4.1% (3.6–4.7%) in 2055, and 0.38% (0.19–0.70%) by 2100 (Figure 1B). Maintaining current testing rates increased testing volume from 17.6 million tests among adults in 2025 to 24.3 million by 2055 and resulted in around 79% of adults living with HIV on ART (Figure 1C). Under this scenario, there were 3.88 (3.37–4.38) million new HIV infections and 1.75 (1.62–1.88) million AIDS-deaths between 2025–2075. This was comparable to the number of new HIV infections that occurred in South Africa in the 13 years from 2009–2022 and AIDS-related deaths in the 16 years from 2006–2022.

**Figure 1:**
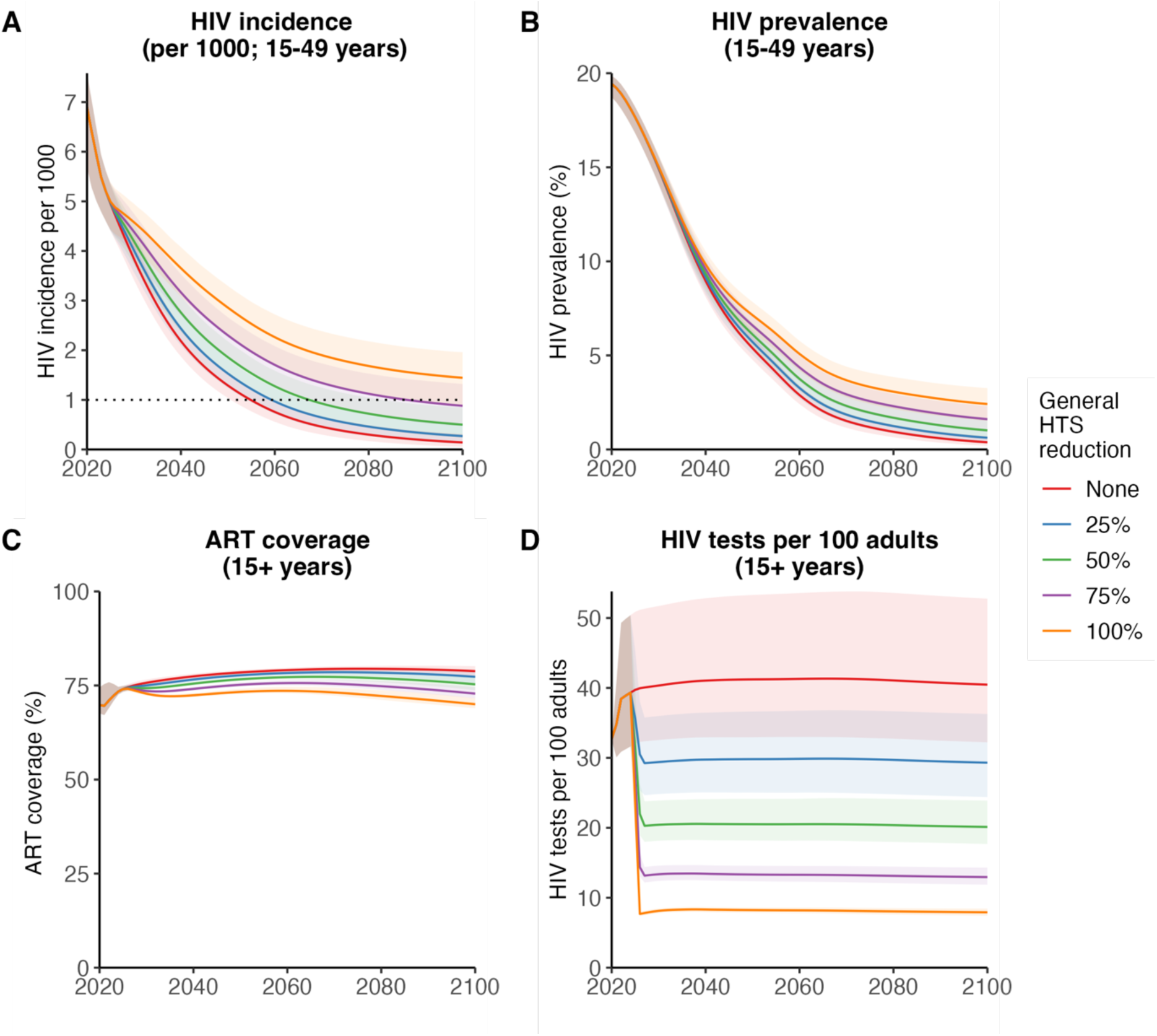
Changes in incidence, prevalence, ART coverage and HIV tests over time when general HTS was reduced. (A) HIV incidence rate (15–49 years) per 1000, (B) HIV prevalence (15–49 years), (C) ART coverage among adult people with HIV (over 15 years), and (D) HIV tests per 100 (over 15 years) between 2020 and 2100. Figures represent status quo scenario (no testing reduction) and general HTS reductions of 25%, 50%, 75% and 100% implemented from 2025. Lines represent posterior mean and shaded areas are 95% confidence intervals. In (A), dotted line represents incidence rate of 1 per 1000 (‘virtual elimination’).

Reducing general HTS in 2025 slowed the decline of HIV incidence, but incidence continued downward even when fully ceasing (100% reduction) general HTS (Figure 1A), retaining only antenatal, symptom-based, and passive partner testing. Fully ceasing general HTS reduced testing volume from around 40 to 8 tests per 100 adults per year (Figure 1D). Reducing general HTS by 25%, 50%, 75% and 100% increased projected HIV incidence in 2100 to 0.27 (0.11–0.53), 0.50 (0.24–0.84), 0.88 (0.47–1.32), and 1.44 (0.86–1.96) per 1000, respectively, between two to ten times higher than 0.14 (0.05–0.31) per 1000 under status quo (Figure 1A). Reducing general HTS by 25%, 50% and 75% delayed incidence <1/1000 by 5, 13, and 35 years beyond 2055 under status quo, and incidence <1/1000 was not attained before 2100 when general HTS was reduced by >80%. HIV prevalence in 2100 was also commensurately higher at 0.62% (0.32–1.07%), 1.02% (0.56–1.59%), 1.61% (0.94–2.31%), and 2.42% (1.53–3.26%) under 25%, 50%, 75% and 100% testing reductions compared to 0.38% (0.19–0.70%) under status quo (Figure 1B).

Scaling-back general HTS reduced long-term ART coverage (Figure 1C) and increased new HIV infections and deaths (Figure S12A-B). ART coverage in 2100 reduced from 78% (77– 80%) in the status quo scenario to 77% (75–79%), 75% (74–77%), 73% (72–75%), and 70% (69–72%), respectively, when scaling-back general HTS by 25% to 100%. This resulted in additional HIV infections between 2025–2075 of 396,000 (299,000–474,000; 10% increase), 926,000 (718,000–1.11 million; 24% increase), 1.63 (1.27–1.96) million (42% increase) and 2.50 (1.97–2.98) million (64% increase). AIDS-deaths increased by 115,000 (94,000–135,000; 7% increase) to 795,000 (670,000–926,000; 45% increase). Despite lower treatment coverage, reducing HTS in 2025 increased the number of adults receiving ART in 2075 from 2.66 (2.34–2.96) million in the status quo scenario to 2.83 (2.48–3.14) million or 3.58 (3.05–3.98) million when reducing general HTS by 25% or 100%, respectively (Figure S12D).

Under status quo, HIV testing was 4.7% (4.0–5.8%) of the total $16.4 billion HIV testing and treatment programme cost over 10 years (2025–2034; undiscounted; Figure S11B). Total costs peaked around 2034 and declined thereafter in all scenarios (Figure S11A), consistent with declining PLWH on ART (Figure S12D). Reducing general HTS by 25% to 100% reduced total costs over 10 years by 1.4% ($236 [177–312] million) to 4.9% ($814 [652– 1032] million; Figure 2B). Most of this reduction was from lower testing costs ($178 [126– 253] million to $541 [412–726] million lower; 75% to 64% of total reductions), while some reduction was from lower ART costs ($72 [59–84] million to $339 [289–385] million lower; 31% to 42% of total reduction) due to fewer PLWH diagnosed and initiated on ART (Figure S12C). Reduced testing lowered annual total costs for around 25 years (between 2025 to 2050), but annual costs were higher after 2050 (Figure 2A) due to more new infections increasing ART and care costs (Figure S11A). Our central estimate suggested that reducing general HTS by 50% would save $180 million (0.27% decrease; Figure 2B) of the total undiscounted cumulative cost over 50 years, however the outcome was uncertain, with 95% CI spanning a $269 million cost increase to $945 million saving. Reducing general HTS by 75% modestly increased the undiscounted cumulative costs by $306 million (0.44% increase), with 95% CI spanning a $629 million cost-saving to a $924 million increase. Ceasing general HTS in 2025 increased 50-year cumulative costs by $1.14 (0.16–1.93) billion (1.59% increase). However, when discounting future costs at 3% or 6% per annum, reducing testing was consistently cost-saving (Figure 2B) because savings occurred in short-term while increased treatment costs were three decades in the future.

**Figure 2:**
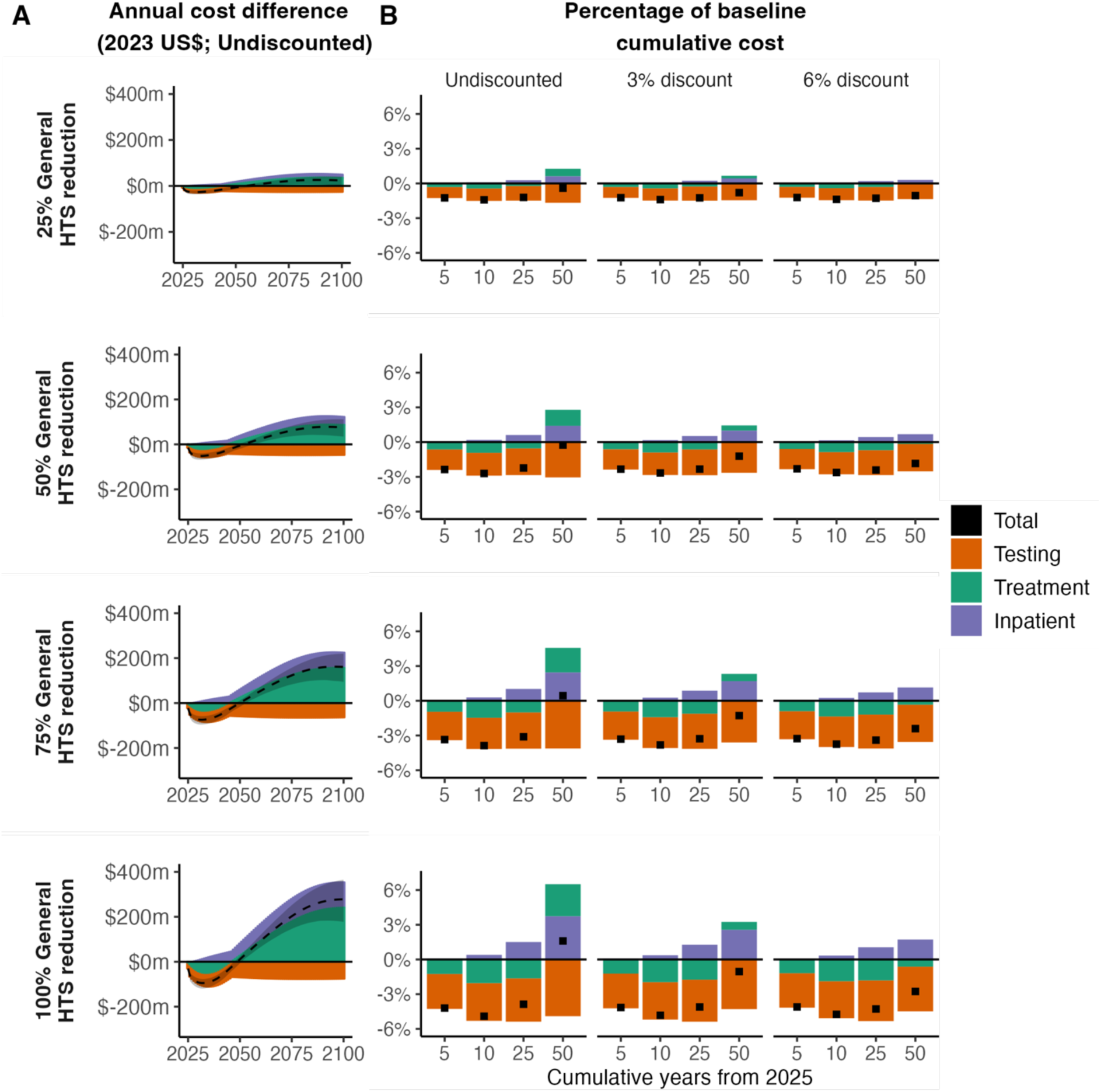
Annual difference from baseline and relative cumulative change from baseline of HIV testing and treatment costs due to general HTS reductions. (A) Annual difference in undiscounted costs from the status quo scenario (baseline) due to general HTS reductions each year between 2020 and 2100 and (B) relative change of the mean cumulative costs due to general HTS reductions as a percentage of the mean cumulative status quo (baseline) cost after 5, 10, 25, and 50 years for 0% (undiscounted), 3%, and 6% discount per annum. The annual cost difference (A) or change from baseline (B) is shown for the total HIV testing and treatment programme (Total), and costs attributable to ANC testing and general HTS (Testing), ART (Treatment), and palliative and inpatient care (Inpatient) when 25%, 50%, 75% and 100% general HTS reductions were implemented from 2025. The negative changes indicate a savings and positive, additional costs due to general HTS reductions. In (A) the dotted line represents the posterior mean and shaded areas are 95% confidence interval for the total costs. All costs are 2023 US$ millions.

Delaying HTS reductions from 2025 until incidence was lower had modest effects on long-term incidence in 2100 (Figure 3A), but larger impacts on the cumulative number of additional infections and deaths (Figure 3B-C). For example, a 50% testing reduction from 2050 resulted in HIV incidence in 2100 of 0.41 (0.17–0.75) per 1000, compared to 0.50 (0.24–0.84) when reduced from 2025. Delaying a 50% testing reduction from 2025 to 2030 decreased new HIV infections from 2025–2075 by 108,000, from 926,000 (718,000–1.11 million) to 818,000 (599,000–998,000; 12% decrease), and additional AIDS-related deaths from 274,000 (225,000–323,000) to 237,000 (192,000–281,000; 13% decrease). Delaying 50% general HTS reduction until 2050 decreased expected new HIV infections by 49% and AIDS-deaths by 54%, however, this still represented 472,000 (268,000– 676,000) additional HIV infections and 126,000 (89,000–163,000) additional AIDS-related deaths from 2050– 2100 compared to if general HTS was not reduced.

**Figure 3:**
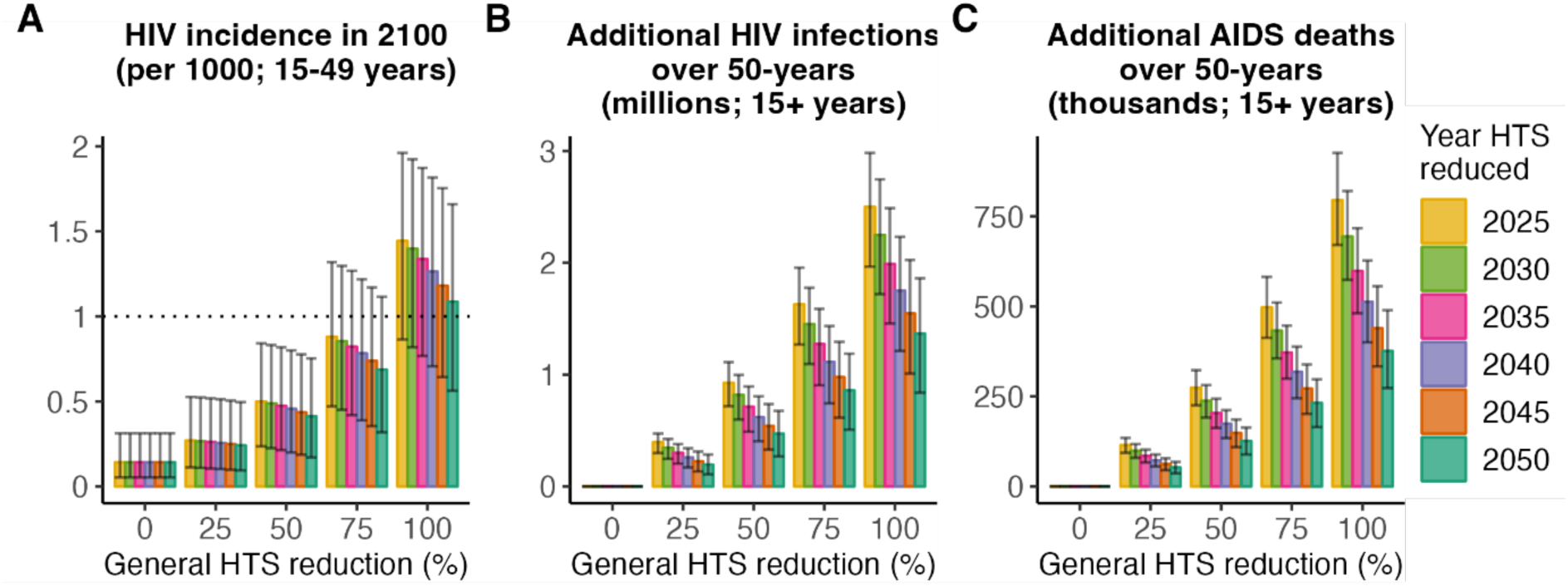
HIV incidence in 2100 and epidemiological costs when general HTS was reduced. Representative figures of mean and 95% CI of HIV incidence (15–49 years) in 2100 (A), additional HIV infections over 50 years (B) and additional AIDS-related deaths over 50 years (C) when testing was reduced by 0, 25%, 50%, 75% and 100% starting from 2025, 2030, 2035, 2040, 2045 or 2050. The dotted line (A) indicates HIV incidence <1/1000 (‘virtual elimination’).

Increasing ART interruption rates between 2025–2035 did not result in rapidly rebounding new HIV infections or deaths (Figure S5), even when combined with reductions in HIV testing (Figure S8), but affected long-term incidence rates, time to <1/1000, and cumulative infections and deaths (Figure 4). With current testing rates but increased ART interruption, incidence still declined steadily if ART coverage reduced to 65% in 2035, but time to incidence <1/1000 was delayed by 15 years to 2070 (2061–2084) (Figure 4B). In the extreme scenario where general HTS ceased from 2025 and ART coverage fell to 50%, incidence in 2100 remained high (4.32 [2.99–5.36] per 1000), comparable to incidence in 2025 (4.95 per 1000; Figure 4A).

**Figure 4.**
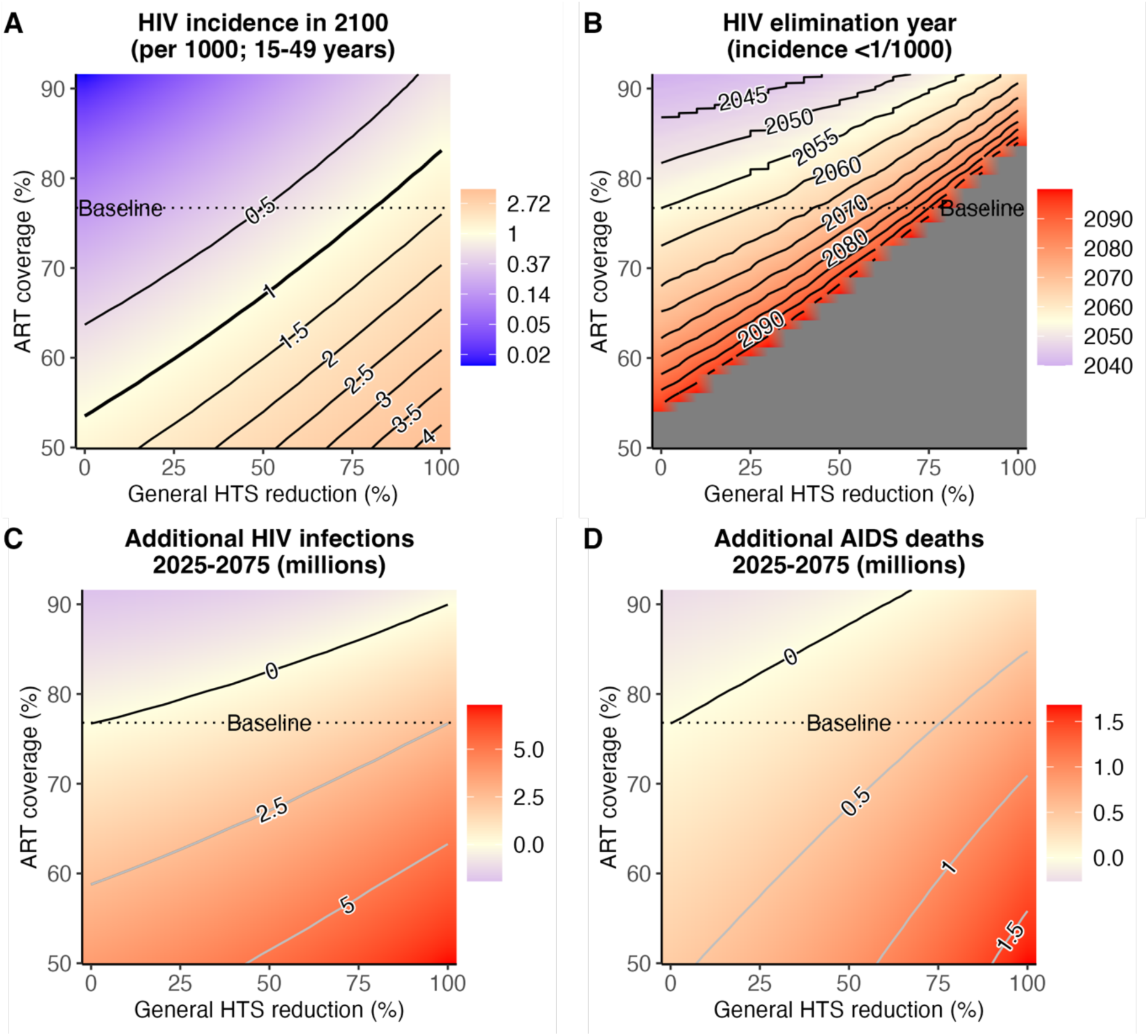
HIV incidence, HIV ‘virtual elimination’ year (incidence <1/1000), additional HIV infections, and AIDS-related deaths when ART coverage was changed in combination with general HTS reductions. Representative heatmaps of HIV incidence per 1000 (15–49 years) in 2100 (A), the year in which HIV incidence <1/1000 (‘virtual elimination’) was attained (B), additional HIV infections over 50 years (C) and additional AIDS-related deaths over 50 years (D) for different combinations of ART coverage in 2035 and testing reductions when testing was reduced from 2025. The colours show the values in the legends (right) for the different combinations – grey (B) indicates combinations where incidence <1/1000 was not attained prior to 2100. The contour lines indicate the incidence in 2100 (A) or incidence <1/1000 year (B) for different combinations, or those combinations for which there are no additional HIV infections (C) or additional AIDS-related deaths (D).

Conversely, improved ART retention could accelerate incidence reductions or enable larger or earlier testing reductions to attain the same epidemiologic impacts. If ART interruption reduced and ART coverage rose to 80%, incidence <1/1000 could be attained by 2055 with up to 25% general HTS reduction, or 75% HTS reduction if ART coverage increased to 90% (Figure 5B). Increasing ART coverage to >90% by 2035 would offset additional new infections caused by ceasing general HTS from 2025 (Figure 4C). However, to avoid additional AIDS-deaths, only a maximum testing reduction of 60% could be borne (Figure 4D). Overall, for every 1% increase in ART coverage in 2035 (up to 90%), general HTS could be reduced by 4.5% in 2025 without increasing expected deaths or infections.

**Figure 5.**
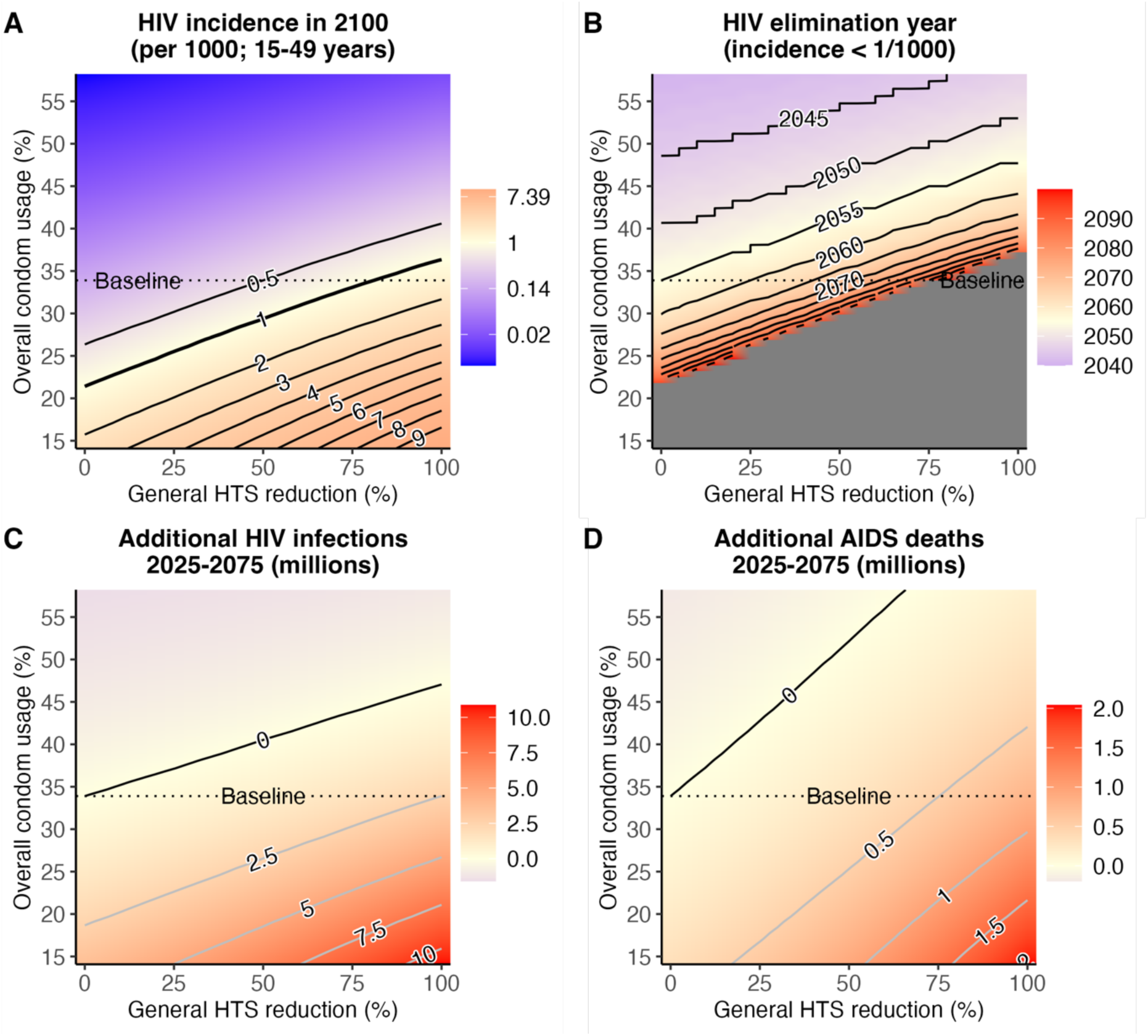
HIV incidence, HIV ‘virtual elimination’ year (incidence <1/1000), additional HIV infections, and AIDS-related deaths when condom usage changes are combined with general HTS reductions. Representative heatmaps of HIV incidence per 1000 (15–49 years) in 2100 (A), the year in which HIV incidence <1/1000 (‘virtual elimination’) was attained (B), additional HIV infections over 50 years (C) and additional AIDS-related deaths over 50 years (D) for different combinations of condom usage in 2035 and testing reductions when testing was reduced from 2025. The colours show the values in the legends (right) for the different combinations – grey (B) indicates combinations where incidence <1/1000 was not attained prior to 2100. The contour lines indicate the incidence in 2100 (A) or incidence <1/1000 year (B) for different combinations, or those combinations for which there are no additional HIV infections (C) or additional AIDS-related deaths (D).

The epidemic trajectory was more sensitive to future changes in sexual risk, such as changes in condom-use. Incidence declined much more slowly if condom-use reduced from 34% to 14% in 2035 and current testing rates were maintained (Figure S7A). Lower future condom-use and reducing general HTS more than 75% reversed declining incidence (Figure 5A, Figure S10A). Any reduction in condom-use delayed attaining incidence <1/1000, with the delay quickly lengthening as risk accumulates. Reducing condom-use from 34% to 26% delayed reaching incidence <1/1000 by 14 years to 2069 (2060–2093), and a fall to 21% made ‘virtual elimination’ unlikely before 2100 (Figure 5B).

Changes in sexual risk could substantially increase infections and deaths over time. Maintaining current testing rates, reduced condom-use to 14% resulted in an additional 3.66 (2.87–4.52) million infections and 346,000 (262,000–442,000) deaths between 2025–2075, compared to the status quo. Conversely, increasing condom usage to 58% by 2035 averted 1.59 (1.29–1.91) million infections and 196,000 (155,000–247,000) deaths. If general HTS were ceased, condom-use falling to 14% led to 10.9 (10.0–11.8) million more infections and 2.04 (1.83–2.21) million more AIDS-deaths, nearly three times more infections and 17% more AIDS-deaths than under status quo. If general HTS ceased in 2025, condom-use would need to increase to >46% to incur the same number of HIV infections as in the status quo (Figure 5C). For every 1% increase in condom usage in 2035 (up to 56%), general HTS could be reduced by 2.7% in 2025 without additional infections or deaths.

## Discussion

Following successful rapid ART coverage increases and progress towards controlling HIV, many countries with large population HIV burdens must consider if, when, and by how much it is reasonable to scale-back resource-intensive programmes, such as general HTS, while avoiding backsliding progress towards reducing new infections and AIDS deaths. Under current conditions, HIV incidence in South Africa is expected to continue steadily declining for several decades, falling to <1/1000 around 2055 and around 1 to 2 per 10,000 by 2100. The proportion of adults aged 15-49 living with HIV will decline from 1 in 5 today to below 1 in 100. We found that managed reductions in HIV testing did not lead to explosive increases in new infections, but slowed the rate of incidence decline and increased the future number of infections, AIDS-related deaths, and requiring lifelong ART. HIV testing contributed only around 5% to total HIV testing and treatment costs. Reducing testing could enable some resources to be allocated to other short-term health system priorities, but at the expense of slower progress towards epidemic control and consequently larger long-term HIV programme costs.

Other countries in eastern and southern Africa with higher ART coverage than South Africa have already begun reducing large-scale general population testing, while promoting more targeted HIV testing approaches, partly due to shifts in donor resources from PEPFAR.^14^ Our findings suggest that these programme shifts are not likely to manifest in reversed progress after one or two decades, but also highlight the long-term cost of slower incidence reductions. Continuing high HIV testing rates for 5 to 25 years longer mitigated some of these impacts. This may be a good strategy, even with low and declining numbers of untreated PLWH, when considering the relatively low cost of HIV testing compared to lifelong HIV care. HIV testing could be increasingly integrated into general disease screening programmes, supporting commitments to universal health coverage.^15,16^ More flexible and targeted HIV testing services for those at higher risk are another strategy to mitigate negative impacts of reducing general HTS.^17^ However, evidence on risk screening tools suggests likely limits to the effectiveness of such strategies.^18^

Our primary projections assumed that ART retention and viral suppression, other HIV prevention (including VMMC, PrEP, condom usage), and sexual risk behaviour are sustained at current levels. In scenarios representing degradation of treatment programme outcomes or increased sexual risk over the next decade, but not catastrophic discontinuation, the combination of increased risk environment and testing reductions did not lead to explosive new infections of the magnitude early in the HIV epidemic or levels unmanageable by the health system. Incidence continued to decline, albeit at a slower rate that could lead to appreciable further HIV infections. In our most extreme scenario, ceasing general HTS in 2025 and reducing condom use from 33% to 14% by 2035 resulted in 11 million more adult infections up to 2075—roughly equivalent to the number who acquired HIV in past 40 years. Our analyses varying ART coverage and condom-use in 2035 within ±20% of their current values represent relatively extreme ranges of plausible changes in a period focused on maintaining coverage of effective interventions; during periods of rapid scale-up, ART coverage (15–49 years) increased from 49% to 70% between 2015 and 2020, while condom use in South Africa is estimated to have increased from 4% to 26% between 1994 and 2004^8,10,19^. These well-managed changes to HIV testing programmes contrast previous modelled scenarios, created to warn against programme disruptions during COVID-19, in which rapid withdrawal of ART from current patients resulted in large immediate increases in infections and AIDS-deaths.^20^

Some changes in programmatic outcomes leading to changes in population risk would be routinely captured, such as ART retention or uptake of prevention. Early detection of decreasing ART coverage could act as triggers for general testing or other interventions to be reimplemented to mitigate accumulation of infection risk. Equally, monitoring improved ART programme outcomes or incidence declines could enable more rapid scaling-back of testing or other intensive interventions without incurring adverse impacts. However, other changes in underlying sexual risk are more difficult to observe and may only be recognised through observing an unexpected change in epidemic trajectory accumulated over a period of 5-10 years or more. This encourages a level of conservatism in HIV programme management decisions. Other outcomes, such as incidence of some sexually transmitted infections, may respond more rapidly to and be useful proxy indicators for changes in the HIV risk environment^21,22^ to enable earlier HIV programme response.

Our study considered the HIV epidemic in South Africa on a longer time horizon than most previous analyses, which have found that HIV incidence in South Africa was not likely to decline below 1 per 1000 up to 2035^23^ or 2063,^9^ while other models have found that Universal Test-and-Treat could theoretically eliminate HIV in South Africa.^24^ The Thembisa model is well-suited for long-term HIV epidemic projections because it incorporates detailed demography, is statistically calibrated to an extensive range of data sources, and incorporates heterogeneity in sexual behaviour, including in partner numbers, engagement in commercial sex work, and sexual mixing. As incidence declines towards elimination, such heterogeneities and key population dynamics become increasingly important in determining epidemic patterns.^9,25,26^ We did not consider optimisation of HIV testing among specific modalities or population groups^7,27^ or the potential impact of scaling-up new prevention strategies such as long-acting injectable antiretrovirals or PrEP^28^ and other interventions optimised in the South Africa HIV Investment Case.^29^ Broadly, effective implementation of these strategies to accelerate declining incidence would affect our conclusions similarly to increased condom-use or increased ART coverage. Our projections are also somewhat more optimistic than recent modelling for eSwatini, which found that incidence declined rapidly as ART coverage reached 90-90-90 targets, followed by steady but slower incidence declines to 2050.^30^

This analysis had several additional limitations. Long-term projections require assumptions about future uncertainties that magnify as the time horizon increases. Our probabilistic analysis represented uncertainties in the parameters driving disease transmission, sexual behaviour, and treatment, and we accounted for potential changes to the HIV programme and sexual risk via scenario analysis. However, our modelling did not consider the full range of factors that could impact the epidemic over the long-term, such future technological advancements (e.g. HIV vaccines or functional cures), other changes in prevention uptake (e.g. VMMC or PrEP), unexpected events that could influence the future of HIV programmes as demonstrated by COVID-19,^20^ or uncertainty about model structure.^31^ Projections of the epidemic to 2100 are therefore more uncertain than the 95% CI may suggest, and should be regarded as a projection of current trends for investigative purposes, rather than a prediction of the future. Long-term HIV programme management decisions are largely driven by considerations of overall cost and efficiency relative to competing public health concerns. Our cost analysis illustrated how programme changes and epidemiologic outcomes translated to short- and long-term HIV programme costs, but we did not quantify the cost-effectiveness compared to other health investment opportunities.

In conclusion, HIV programmes with high testing rates designed to reach high population HIV burden can likely scale-back general HTS without risking an immediate explosion of new infections or AIDS deaths. Testing costs are small compared to ART provision, so reducing testing only modestly saved short-term costs and likely increased long-term costs due to more HIV infections, AIDS deaths, and PLWH requiring ART. Therefore, HIV programmes face policy decisions about the opportunity to save or reallocate short-term resources by reducing testing, balanced with how rapidly to continue progressing towards reducing infections and controlling HIV. Changes in testing had a slow impact on incidence trends, but reduced testing combined with changes to the sexual risk environment could derail progress towards controlling HIV if programmatic changes happen too early, highlighting the need for caution in making large programmatic changes and continued monitoring to adjust programmes.

## Data Availability

All data produced in the present work are contained in the manuscript

## Author contributions

The study was conceived by SPR, LFJ, and JWI-E with input from LKW and GM-R. The model was designed by LFJ, and implemented for this study by LFJ and SPR. Analysis was performed by SPR. GM-R and LJ developed the costing model. LFJ acquired study funding. All authors had access to the data and accept responsibility for submission for publication. The report was drafted by SPR and LKW and revised for intellectual content by all authors.

## Supplementary Materials

### Assumptions and calibration of testing, ART, and condom-use in Thembisa model

In the Thembisa model, testing rates are calibrated to national data on total test numbers, test positivity, and the proportion of adults reporting having ever tested.^32^ Following a positive test, ART initiation rates depend on CD4 count and sex, and vary over time calibrated to ART programme scale-up. People on ART interrupt treatment at a time-constant rate that differs by sex but is independent of age, CD4 count, and ART duration. The female interruption rate was estimated during model calibration. The male rate was 1.2 times the female rate, based on previous studies.^33^ The rate of resuming ART after interruption varied over time according to population HIV testing rates and by sex, with women resuming ART 1.5 times more rapidly than men. Modelled probability of condom-use differs by relationship type, age, sex, knowledge of HIV status and ART use among PLWH, and incorporates temporal trends estimated from statistical analysis of nationally representative survey data (1986-2017).^34^

### Projection assumptions about male circumcision and pre-exposure prophylaxis

Thembisa model projections incorporated continued provision of voluntary medical male circumcision (VMMC) and pre-exposure prophylaxis (PrEP) among higher risk populations. Male circumcision is assumed to lower male HIV acquisition by 60%, and has no effect on male-to-female or male-to-male HIV transmission rates. Circumcision rates represent both background traditional male circumcision, calibrated to circumcision data prior to 2008,^35^ and VMMC implemented for HIV prevention. From 2021 onwards, the annual probability of VMMC in boys aged 10–14 years is assumed to be 0.26 per year and decreased with age to 0.15, 0.07, 0.036, and 0.003 per year among age groups 15–19, 20–24, 25–49, and 50+ years, respectively. Under these assumptions, total circumcision coverage among men 15-49 years increased from 61% in 2021 to 91% in 2060 and was stable thereafter.

PrEP rollout was initiated in 2016 among FSW and among MSM in 2017. This was expanded to university students in 2018, adolescents and young women (AGYW) (15–24 years) from 2019, and the general population from 2021. In all population groups besides FSW, PrEP uptake is assumed to be proportional to the level of HIV risk, quantified by the product of the annual number of sexual partners (determined by age, risk group and marital status) and the HIV prevalence of those partners (determined by sex and key population status). From 2021–2025 the monthly PrEP initiation rate among PrEP eligible FSW is assumed to increase from 4.2% to 6.0% and remain at that rate until 2100. PrEP initiation rates among high-risk MSM aged 20 years and high-risk women aged 20 years who are PrEP eligible was assumed to be half that of FSW, increasing from 2.1% to 3.0% per month over 2021–2025. PrEP efficacy was assumed to be 65% in heterosexuals^36^ and 85% in MSM.^37,38^ Average duration of PrEP continuation was assumed to be 6-months among FSW and AGYW and one year among MSM. Under these PrEP initiation and continuation assumptions, the percentage of MSM on PrEP increased from 14% in 2021 to 42% in 2100 while the percentage of FSW on PrEP increased from 22% in 2021 to 34% in 2026 and was stable thereafter, and AGYW on PrEP increased from 5% in 2021 to 9% in 2026 and was stable thereafter.

### Changes to Thembisa model to implement long-term projection scenarios

There were two changes made to the Thembisa 4.5 model. Firstly, to extend Thembisa projections year to 2100, we changed the maximum projection years from 86 (1985 to 2070) to 116 (1985 to 2100). Annual rates and proportions used in the input parameters were extended to 2100 assuming continuation of the same values that were constant between 2021 and 2070 up to 2100, including demographic rate parameters, time-varying HIV risk parameters (e.g. condom usage proportions), rates of HIV testing, care, and treatment, and uptake of male circumcision and PrEP for HIV prevention.

Secondly, in Thembisa 4.5, condom usage is defined as a probability for the three different partnership types, namely short-term (ST), long-term (LT), and female sex worker (FSW)-client. To change the condom usage for all three relationships by the same relative value while preventing the number of sex acts in which condom are used from exceeding the total sex acts, we defined an odds ratio for condom usage by relationship. In scenarios varying condom use probabilities, the probability of condom usage for each relationship was converted to the odds-scale, adjusted by the odds-ratio, and then returned to the probability scale.

### Modelled scenarios

Tables S1–S4 summarise the scenarios that were performed for the analysis.

**Table S1.** Baseline.

| Testing rate | Testing reduction year | ART interruption rate | Condom usage odds |
| --- | --- | --- | --- |
| 2021 base rate | N/A | 2021 base rate | 2021 base odds |

**Table S2.**
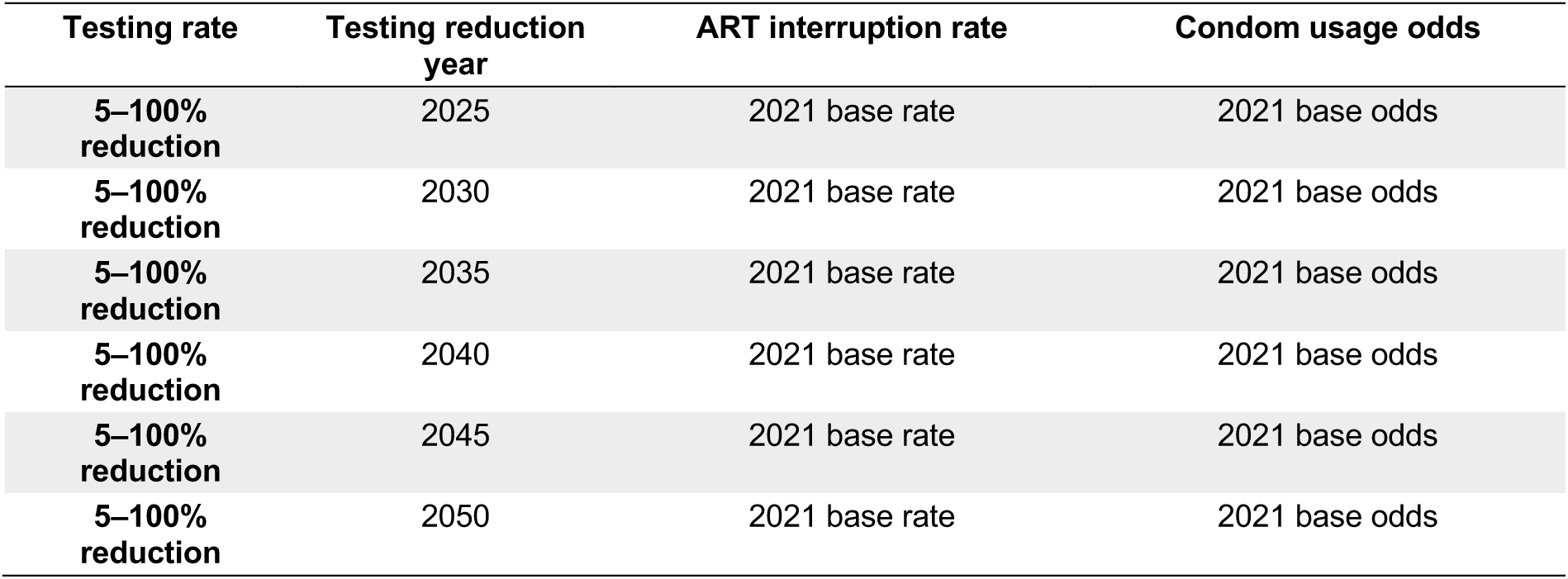
Testing reductions with status quo ART coverage and condom usage.

**Table S3.** Testing reductions with altered ART coverage and condom usage.

| Testing rate | Testing reduction year | ART interruption rate | Condom usage odds |
| --- | --- | --- | --- |
| 2021 base rate | N/A | 0.5–14% increase over 10 years from 2025 | 2021 base odds |
| 2021 base rate | N/A | 0.5–14% decrease over 10 years from 2025 | 2021 base odds |
| 5–100% reduction | 2025 | 0.5–14% increase over 10 years from 2025 | 2021 base odds |
| 5–100% reduction | 2025 | 0.5–14% decrease over 10 years from 2025 | 2021 base odds |
| 5–100% reduction | 2030 | 0.5–14% increase over 10 years from 2025 | 2021 base odds |
| 5–100% reduction | 2030 | 0.5–14% decrease over 10 years from 2025 | 2021 base odds |
| 5–100% reduction | 2035 | 0.5–14% increase over 10 years from 2025 | 2021 base odds |
| 5–100% reduction | 2035 | 0.5–14% decrease over 10 years from 2025 | 2021 base odds |
| 5–100% reduction | 2040 | 0.5–14% increase over 10 years from 2025 | 2021 base odds |
| 5–100% reduction | 2040 | 0.5–14% decrease over 10 years from 2025 | 2021 base odds |
| **5–100%**<br>**reduction** | 2045 | 0.5–14% increase over 10 years from 2025 | 2021 base odds |
| **5–100%**<br>**reduction** | 2045 | 0.5–14% decrease over 10 years from 2025 | 2021 base odds |
| **5–100%**<br>**reduction** | 2050 | 0.5–14% increase over 10 years from 2025 | 2021 base odds |
| **5–100%**<br>**reduction** | 2050 | 0.5–14% decrease over 10 years from 2025 | 2021 base odds |

### Cost analysis assumptions

The cost analysis was performed by multiplying the units by the average cost per unit in Table S5. Assumptions on outputs used in the cost analysis are shown in Table S6. Costs were estimated from the providers’ perspective (South African government) and presented in 2023 United States dollar (US$). Costs are calculated as fully-loaded costs, including the cost of staff time, consumables, lab tests (if applicable), drugs (if applicable), equipment, and overheads (updated based on previous analysis^39^).

**Table S4.**
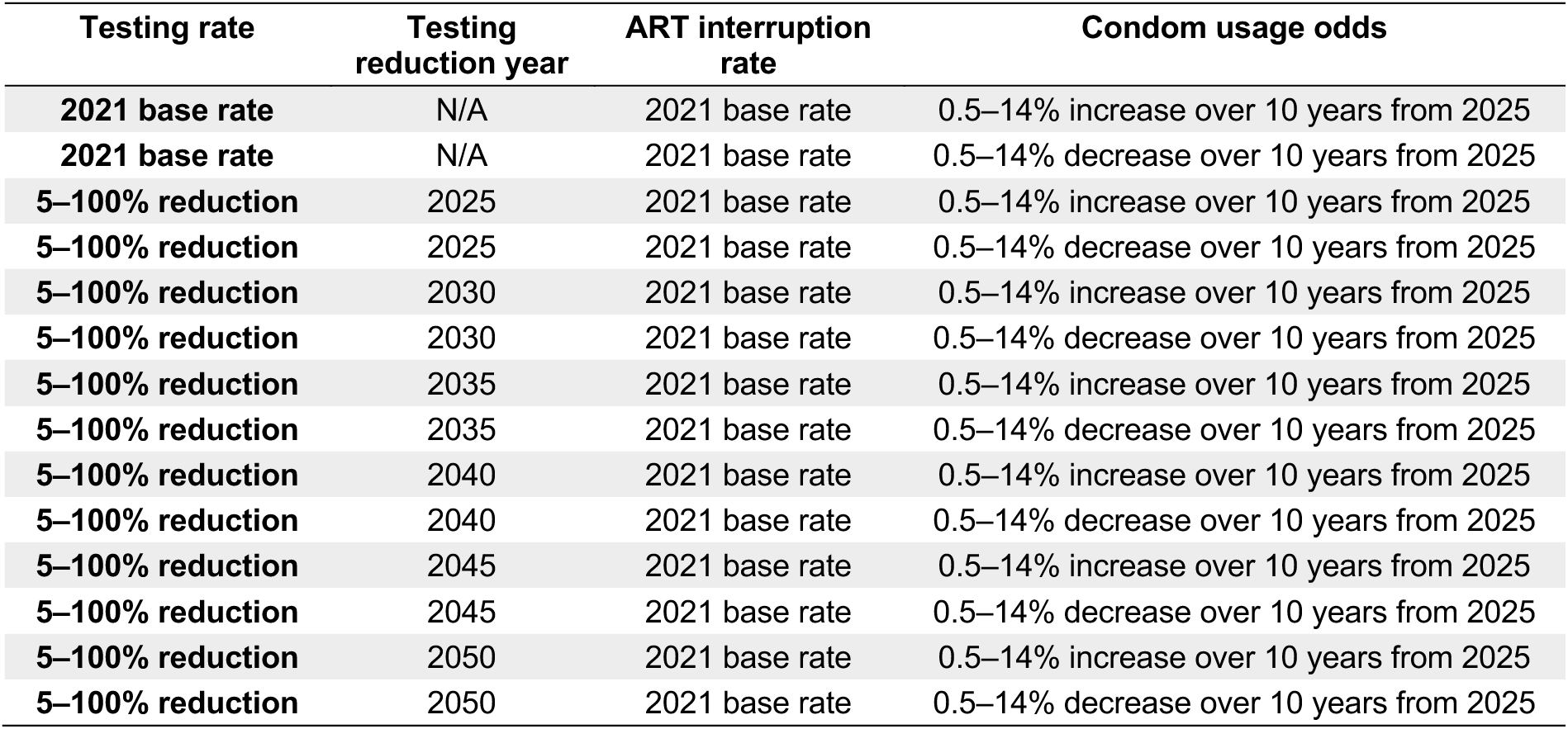
Testing reductions with status quo ART coverage and altered condom usage.

| Testing rate | Testing reduction year | ART interruption rate | Condom usage odds |
| --- | --- | --- | --- |
| **2021 base rate** | N/A | 2021 base rate | 0.5–14% increase over 10 years from 2025 |
| **2021 base rate** | N/A | 2021 base rate | 0.5–14% decrease over 10 years from 2025 |
| **5–100% reduction** | 2025 | 2021 base rate | 0.5–14% increase over 10 years from 2025 |
| **5–100% reduction** | 2025 | 2021 base rate | 0.5–14% decrease over 10 years from 2025 |
| **5–100% reduction** | 2030 | 2021 base rate | 0.5–14% increase over 10 years from 2025 |
| **5–100% reduction** | 2030 | 2021 base rate | 0.5–14% decrease over 10 years from 2025 |
| **5–100% reduction** | 2035 | 2021 base rate | 0.5–14% increase over 10 years from 2025 |
| **5–100% reduction** | 2035 | 2021 base rate | 0.5–14% decrease over 10 years from 2025 |
| **5–100% reduction** | 2040 | 2021 base rate | 0.5–14% increase over 10 years from 2025 |
| **5–100% reduction** | 2040 | 2021 base rate | 0.5–14% decrease over 10 years from 2025 |
| **5–100% reduction** | 2045 | 2021 base rate | 0.5–14% increase over 10 years from 2025 |
| **5–100% reduction** | 2045 | 2021 base rate | 0.5–14% decrease over 10 years from 2025 |
| **5–100% reduction** | 2050 | 2021 base rate | 0.5–14% increase over 10 years from 2025 |
| **5–100% reduction** | 2050 | 2021 base rate | 0.5–14% decrease over 10 years from 2025 |

**Table S5.**
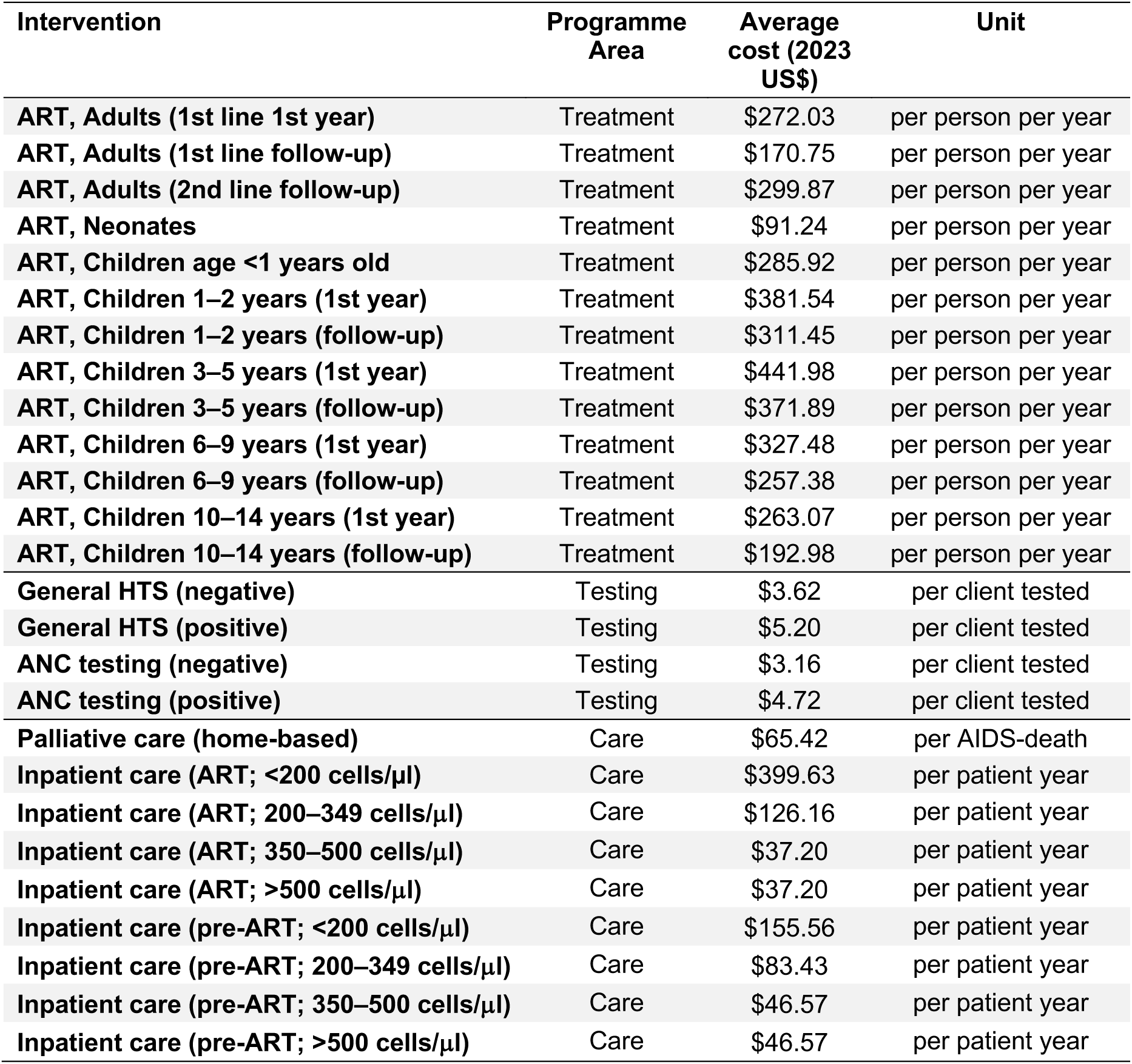
Costs per unit for different intervention in SA HIV programme.

**Table S6.**
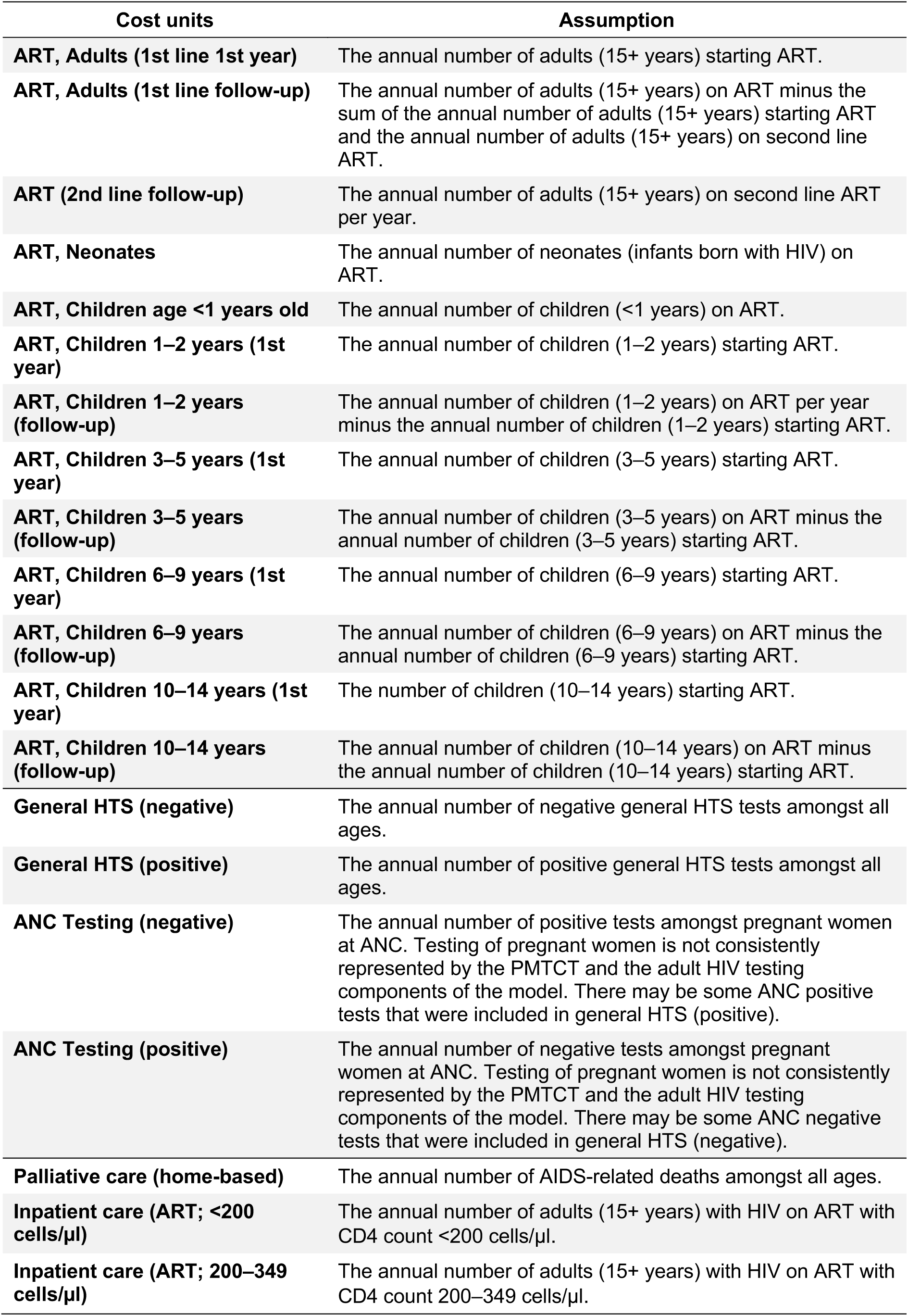
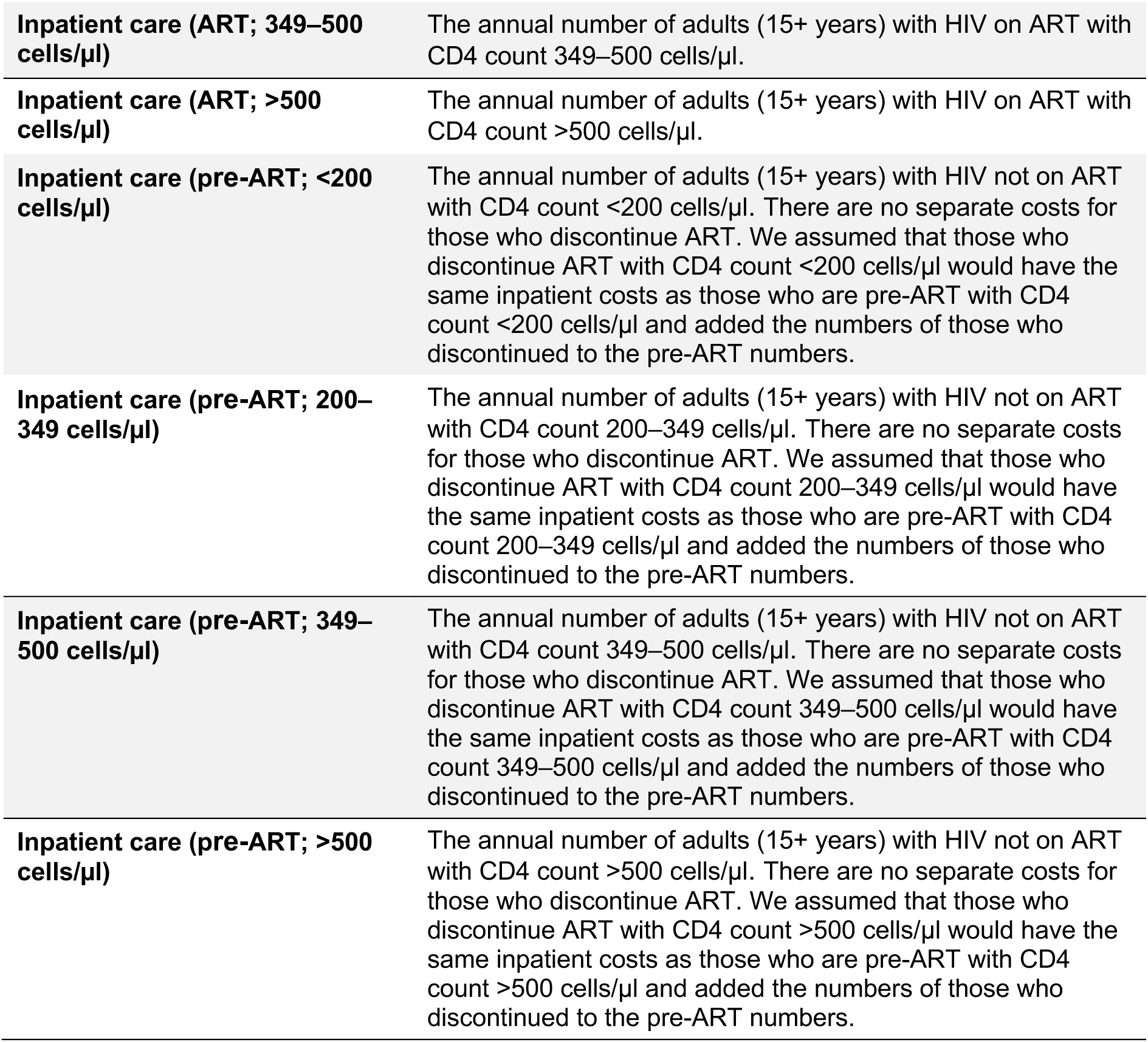
Assumptions applied to cost analysis.

| Cost units | Assumption |
| --- | --- |
| **ART, Adults (1st line 1st year)** | The annual number of adults (15+ years) starting ART. |
| **ART, Adults (1st line follow-up)** | The annual number of adults (15+ years) on ART minus the sum of the annual number of adults (15+ years) starting ART and the annual number of adults (15+ years) on second line ART. |
| **ART (2nd line follow-up)** | The annual number of adults (15+ years) on second line ART per year. |
| **ART, Neonates** | The annual number of neonates (infants born with HIV) on ART. |
| **ART, Children age <1 years old** | The annual number of children (<1 years) on ART. |
| **ART, Children 1–2 years (1st year)** | The annual number of children (1–2 years) starting ART. |
| **ART, Children 1–2 years (follow-up)** | The annual number of children (1–2 years) on ART per year minus the annual number of children (1–2 years) starting ART. |
| **ART, Children 3–5 years (1st year)** | The annual number of children (3–5 years) starting ART. |
| **ART, Children 3–5 years (follow-up)** | The annual number of children (3–5 years) on ART minus the annual number of children (3–5 years) starting ART. |
| **ART, Children 6–9 years (1st year)** | The annual number of children (6–9 years) starting ART. |
| **ART, Children 6–9 years (follow-up)** | The annual number of children (6–9 years) on ART minus the annual number of children (6–9 years) starting ART. |
| **ART, Children 10–14 years (1st year)** | The number of children (10–14 years) starting ART. |
| **ART, Children 10–14 years (follow-up)** | The annual number of children (10–14 years) on ART minus the annual number of children (10–14 years) starting ART. |
| **General HTS (negative)** | The annual number of negative general HTS tests amongst all ages. |
| **General HTS (positive)** | The annual number of positive general HTS tests amongst all ages. |
| **ANC Testing (negative)** | The annual number of positive tests amongst pregnant women at ANC. Testing of pregnant women is not consistently represented by the PMTCT and the adult HIV testing components of the model. There may be some ANC positive tests that were included in general HTS (positive). |
| **ANC Testing (positive)** | The annual number of negative tests amongst pregnant women at ANC. Testing of pregnant women is not consistently represented by the PMTCT and the adult HIV testing components of the model. There may be some ANC negative tests that were included in general HTS (negative). |
| **Palliative care (home-based)** | The annual number of AIDS-related deaths amongst all ages. |
| **Inpatient care (ART; <200 cells/μl)** | The annual number of adults (15+ years) with HIV on ART with CD4 count <200 cells/μl. |
| **Inpatient care (ART; 200–349 cells/μl)** | The annual number of adults (15+ years) with HIV on ART with CD4 count 200–349 cells/μl. |
| **Inpatient care (ART; 349–500 cells/μl)** | The annual number of adults (15+ years) with HIV on ART with CD4 count 349–500 cells/μl. |
| **Inpatient care (ART; >500 cells/μl)** | The annual number of adults (15+ years) with HIV on ART with CD4 count >500 cells/μl. |
| **Inpatient care (pre-ART; <200 cells/μl)** | The annual number of adults (15+ years) with HIV not on ART with CD4 count <200 cells/μl. There are no separate costs for those who discontinue ART. We assumed that those who discontinue ART with CD4 count <200 cells/μl would have the same inpatient costs as those who are pre-ART with CD4 count <200 cells/μl and added the numbers of those who discontinued to the pre-ART numbers. |
| **Inpatient care (pre-ART; 200–349 cells/μl)** | The annual number of adults (15+ years) with HIV not on ART with CD4 count 200–349 cells/μl. There are no separate costs for those who discontinue ART. We assumed that those who discontinue ART with CD4 count 200–349 cells/μl would have the same inpatient costs as those who are pre-ART with CD4 count 200–349 cells/μl and added the numbers of those who discontinued to the pre-ART numbers. |
| **Inpatient care (pre-ART; 349–500 cells/μl)** | The annual number of adults (15+ years) with HIV not on ART with CD4 count 349–500 cells/μl. There are no separate costs for those who discontinue ART. We assumed that those who discontinue ART with CD4 count 349–500 cells/μl would have the same inpatient costs as those who are pre-ART with CD4 count 349–500 cells/μl and added the numbers of those who discontinued to the pre-ART numbers. |
| **Inpatient care (pre-ART; >500 cells/μl)** | The annual number of adults (15+ years) with HIV not on ART with CD4 count >500 cells/μl. There are no separate costs for those who discontinue ART. We assumed that those who discontinue ART with CD4 count >500 cells/μl would have the same inpatient costs as those who are pre-ART with CD4 count >500 cells/μl and added the numbers of those who discontinued to the pre-ART numbers. |

**Figure S1:**
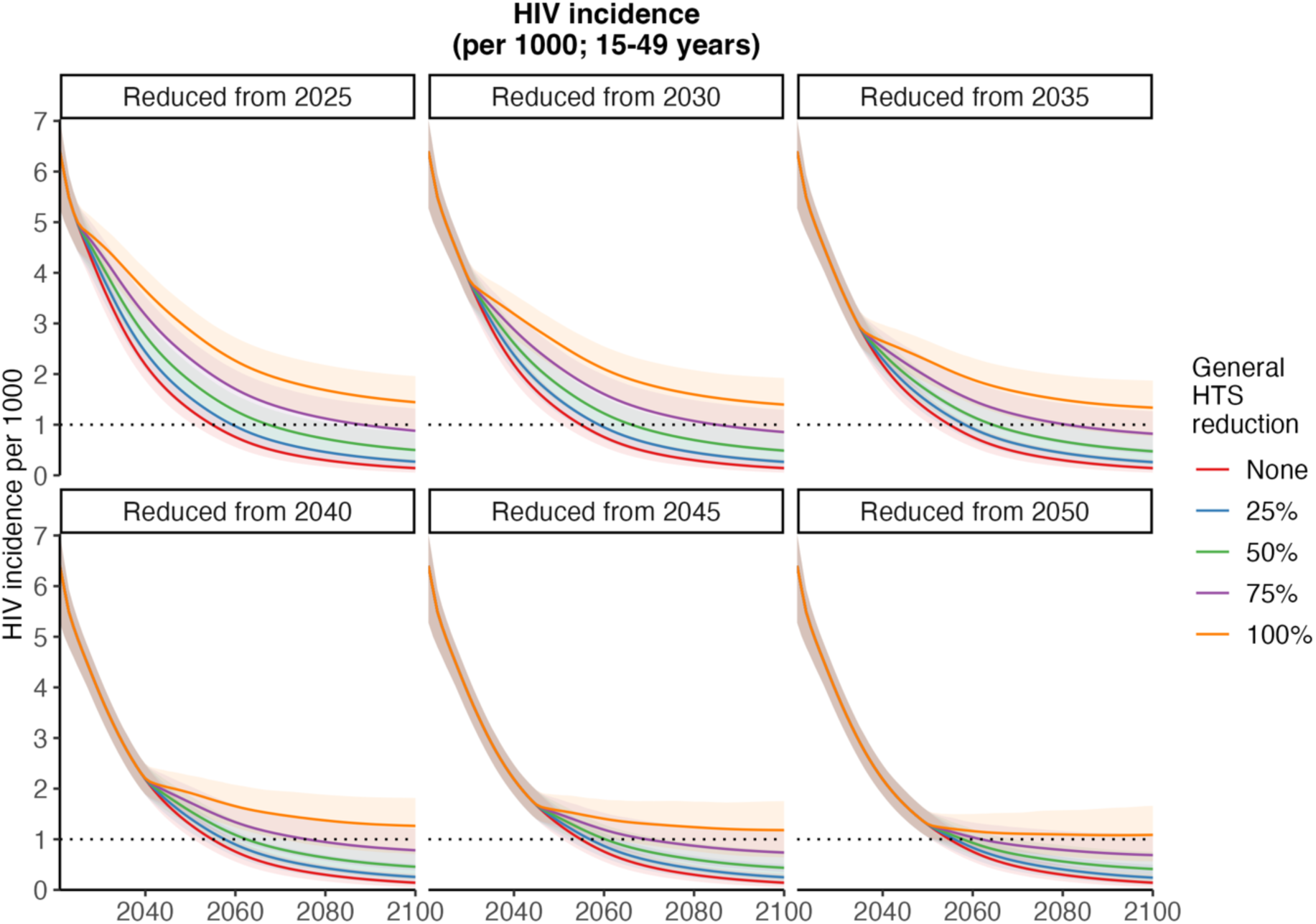
Sensitivity of HIV incidence to timing of general HTS reductions. Changes in incidence over time when general HTS was reduced from different time points. Figures of HIV incidence rate (15–49 years) per 1000 (mean and 95% CI) including an indication of when ‘elimination’ was attained (dotted line) between 2020 and 2100. Lines show status quo (no testing reduction) and general HTS reductions of 25%, 50%, 75% and 100% when testing was reduced from 2025, 2030, 2035, 2040, 2045 and 2050.

**Figure S2:**
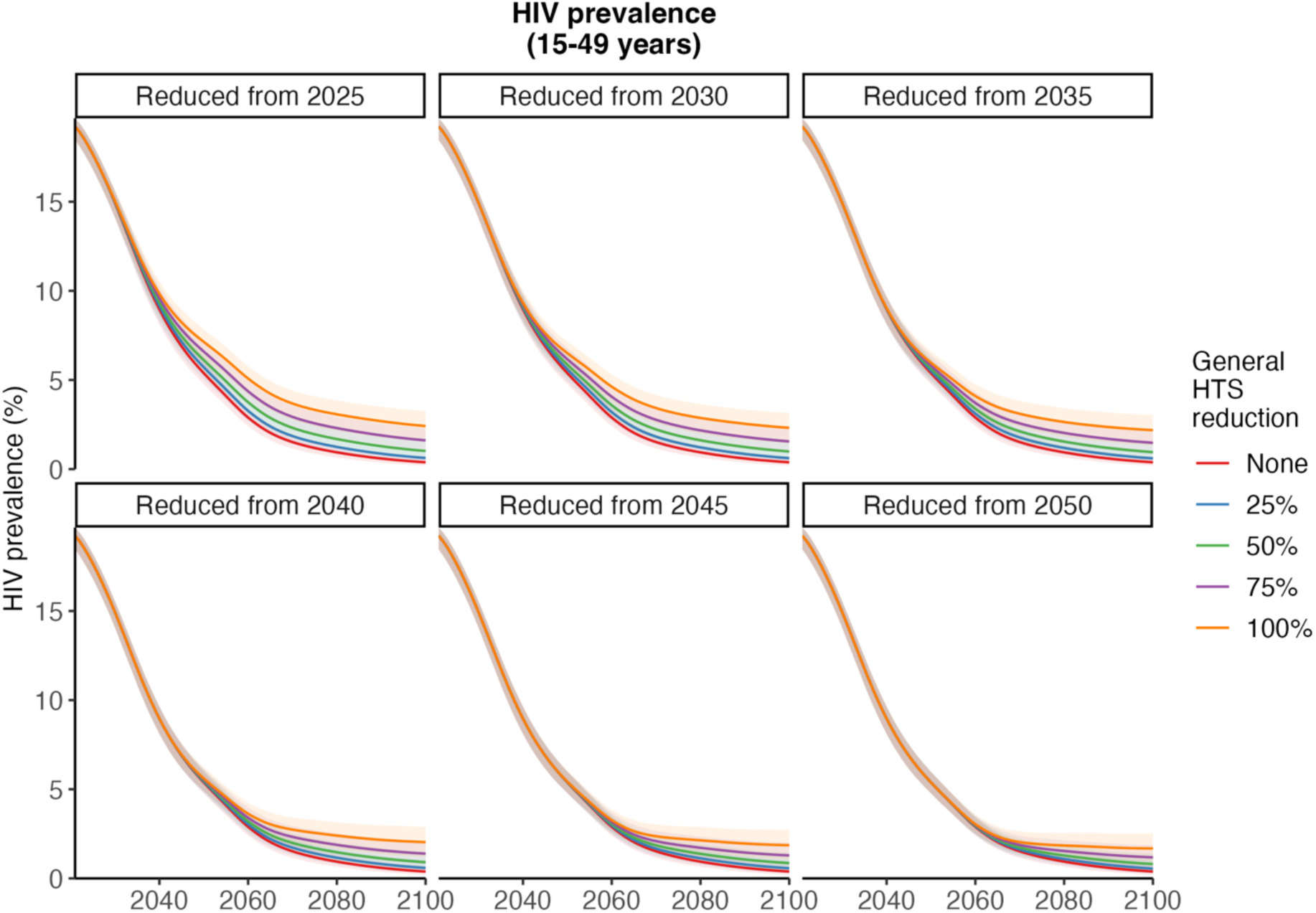
Sensitivity of HIV prevalence to timing of general HTS reductions. Changes in HIV prevalence when general HTS was reduced from different time points. Figures of HIV prevalence (%) (15–49 years) (mean and 95% CI) between 2020 and 2100. Lines show status quo (no testing reduction) and general HTS reductions of 25%, 50%, 75% and 100% when testing was reduced from 2025, 2030, 2035, 2040, 2045 and 2050.

**Figure S3:**
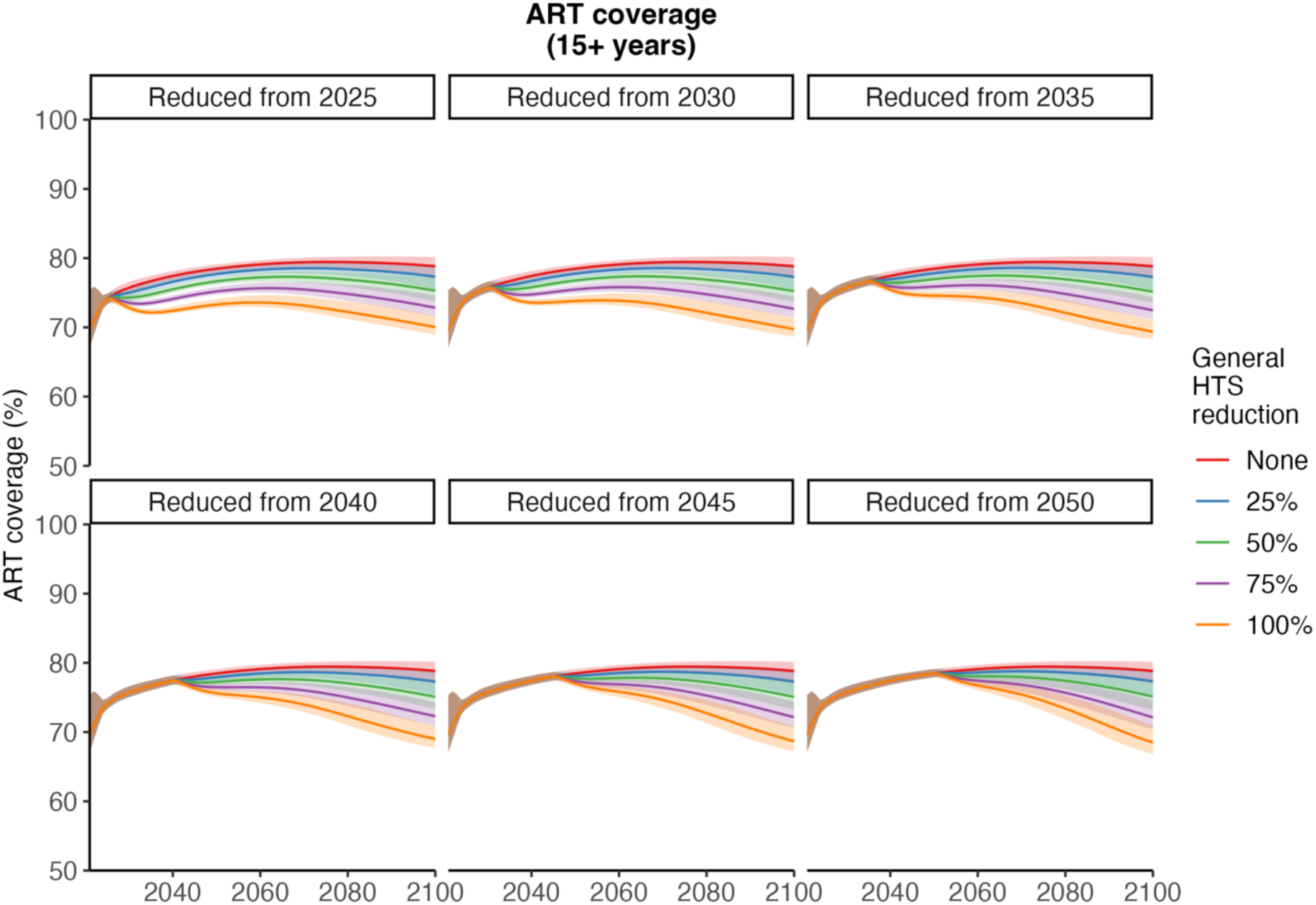
Sensitivity of ART coverage to timing of general HTS reductions. Changes in ART coverage when general HTS was reduced from different time points. Figures of ART coverage (%) (over 15 years) (mean and 95% CI) between 2020 and 2100. Lines show status quo (no testing reduction) and general HTS reductions of 25%, 50%, 75% and 100% when testing was reduced from 2025, 2030, 2035, 2040, 2045 and 2050.

**Figure S4:**
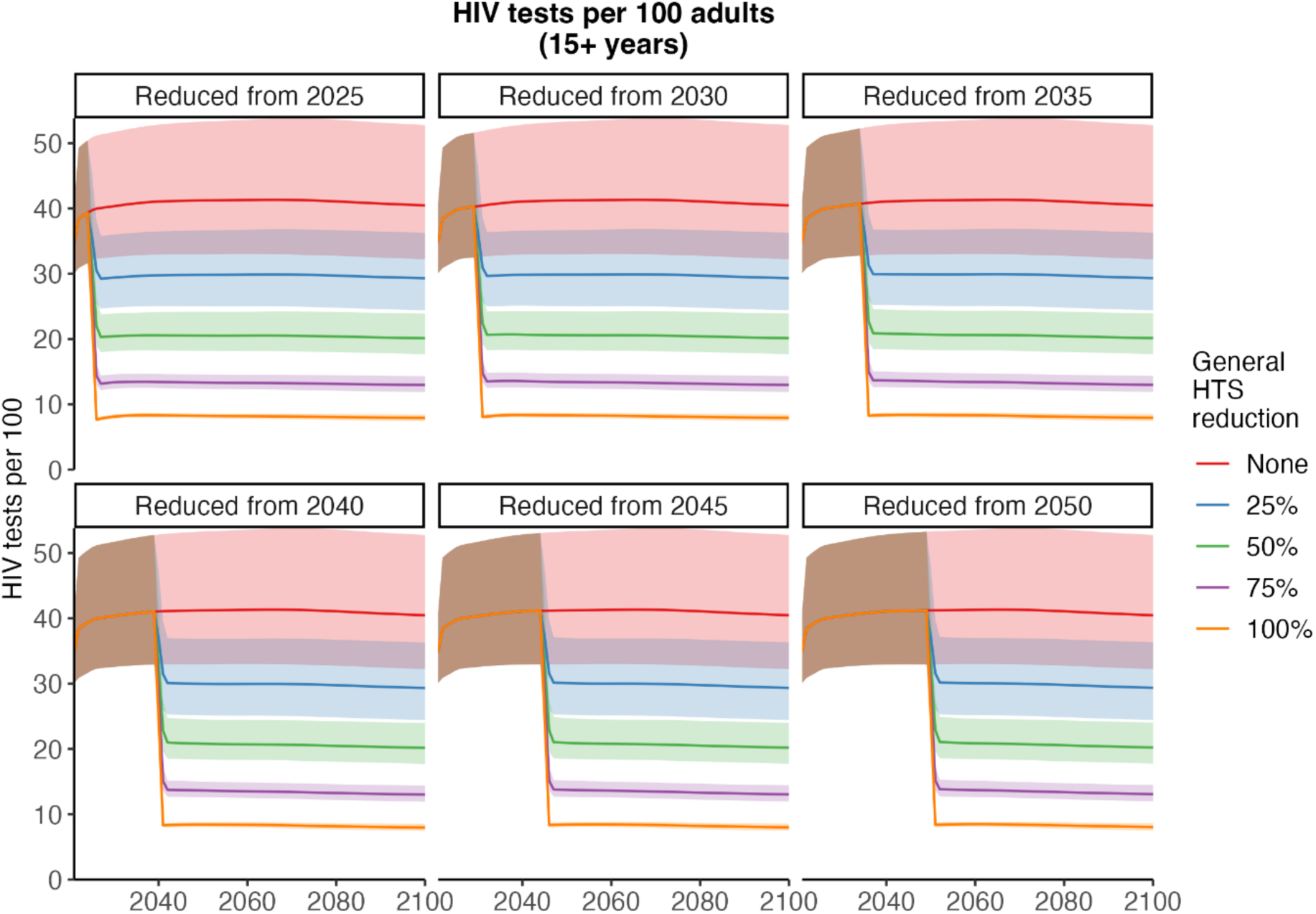
Sensitivity of HIV testing to timing of general HTS reductions. Changes in HIV tests when general HTS was reduced from different time points. Figures of HIV tests per 1000 adults (over 15 years) (mean and 95% CI) between 2020 and 2100. Lines show status quo (no testing reduction) and general HTS reductions of 25%, 50%, 75% and 100% when testing was reduced from 2025, 2030, 2035, 2040, 2045 and 2050.

**Figure S5:**
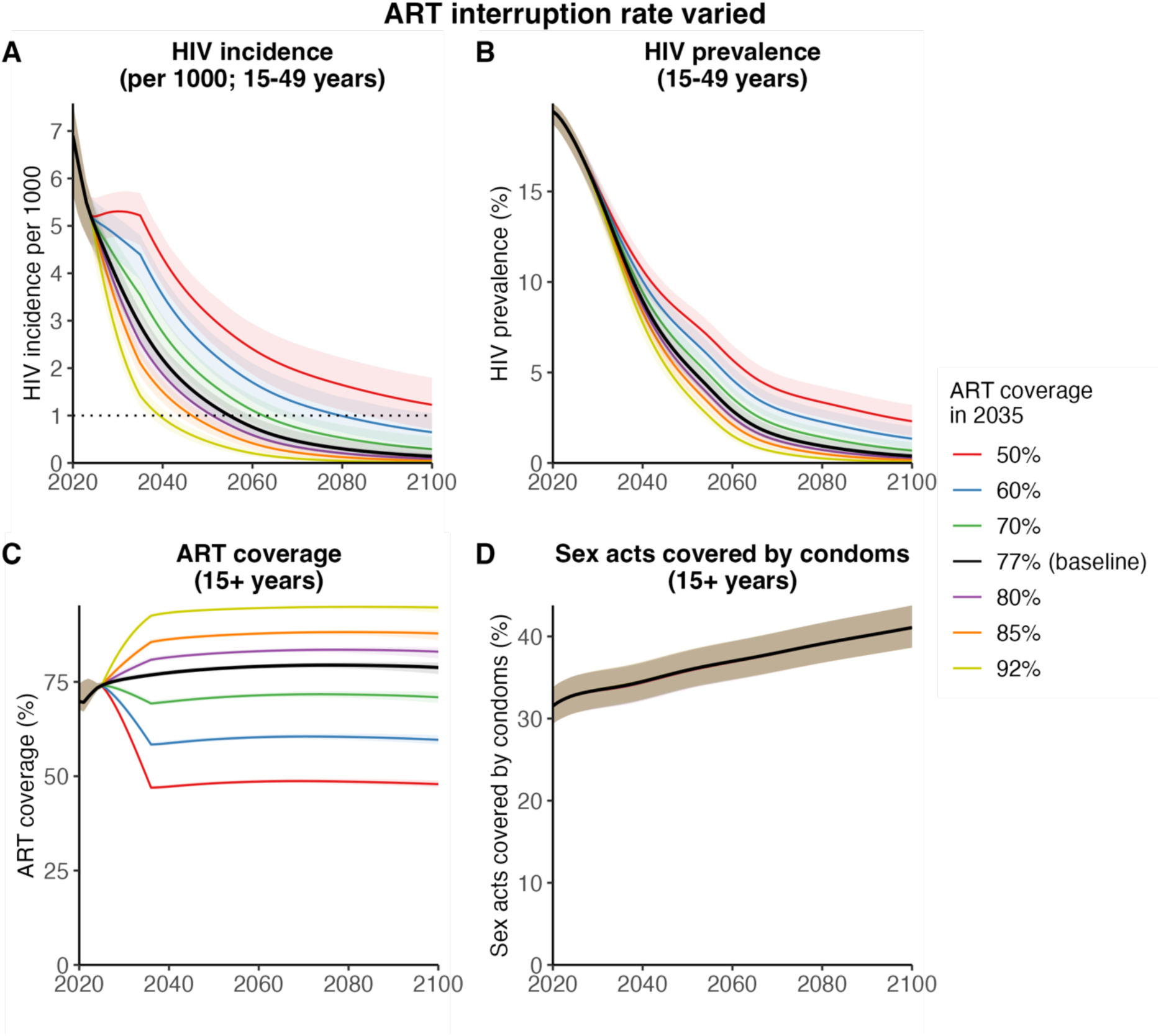
Sensitivity of HIV incidence and prevalence to future changes in ART interruption rate. Changes in incidence, prevalence, ART coverage and condom usage over time when ART coverage was increased or decreased. Representative figures of HIV incidence rate (15–49 years) (A) per 1000 (mean and 95% CI) including an indication of when ‘virtual elimination’ (incidence <1/1000) was attained (dotted line), HIV prevalence (15–49 years) (B), ART coverage of adults (over 15 years) (C) and condom usage (%) (over 15 years) (D) between 2020 and 2100 showing status quo testing when ART coverage in 2035 was either retained at baseline (77%), increased to 80%, 85% or 92%, or decreased to 70%, 60%, or 50%.

**Figure S7:**
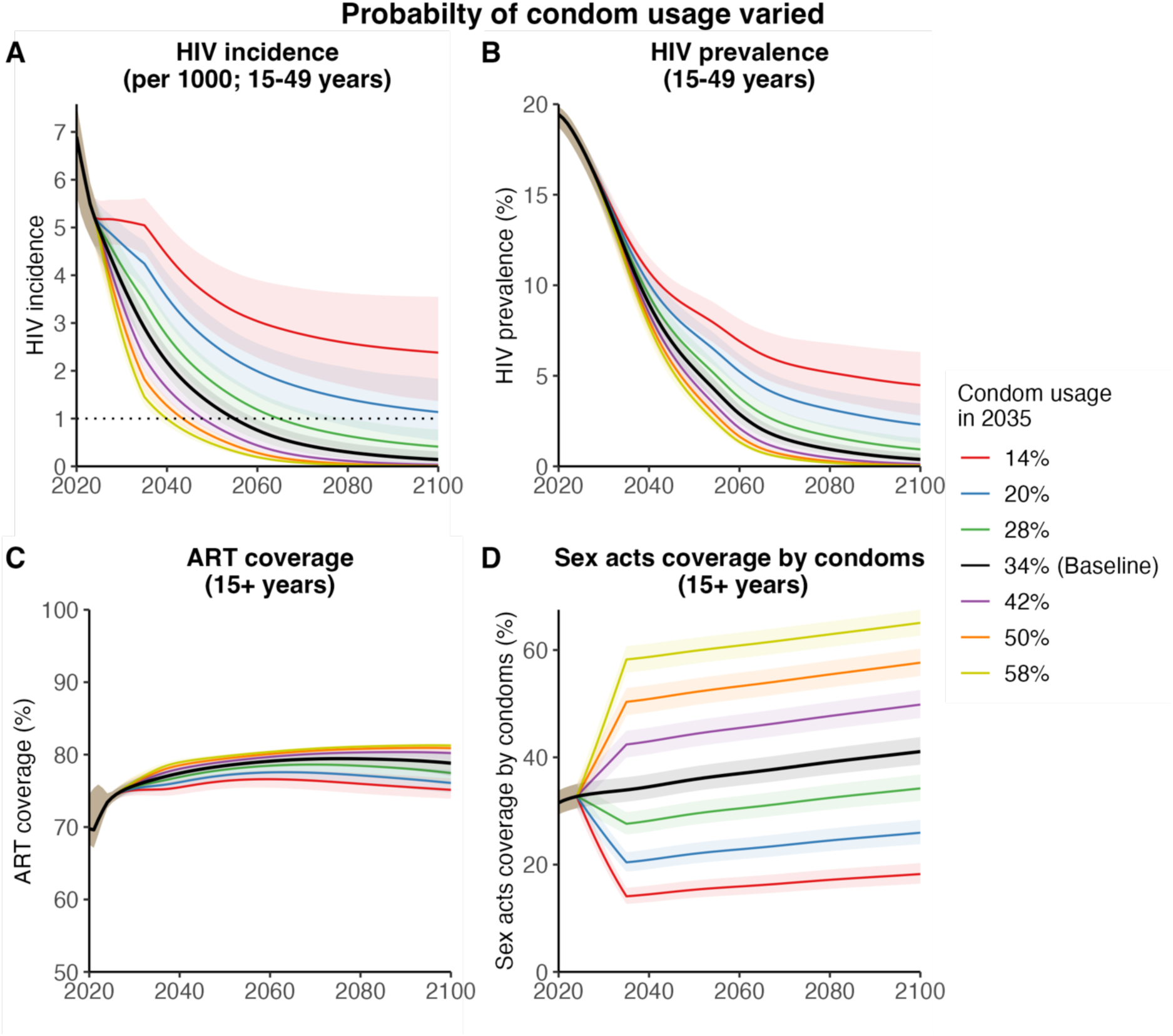
Sensitivity of HIV incidence and prevalence to future changes in probability of condom usage. Changes in incidence, prevalence, ART coverage and condom usage over time when ART coverage was increased or decreased. Representative figures of HIV incidence rate (15– 49 years) (A) per 1000 (mean and 95% CI) including an indication of when ‘virtual elimination’ (incidence <1/1000) was attained (dotted line), HIV prevalence (15–49 years) (B), ART coverage of adults (over 15 years) (C) and condom usage (%) (over 15 years) (D) between 2020 and 2100 showing status quo testing when ART coverage in 2035 was either retained at baseline (77%), increased to 80%, 85% or 92%, or decreased to 70%, 60%, or 50%.

**Figure S7:**
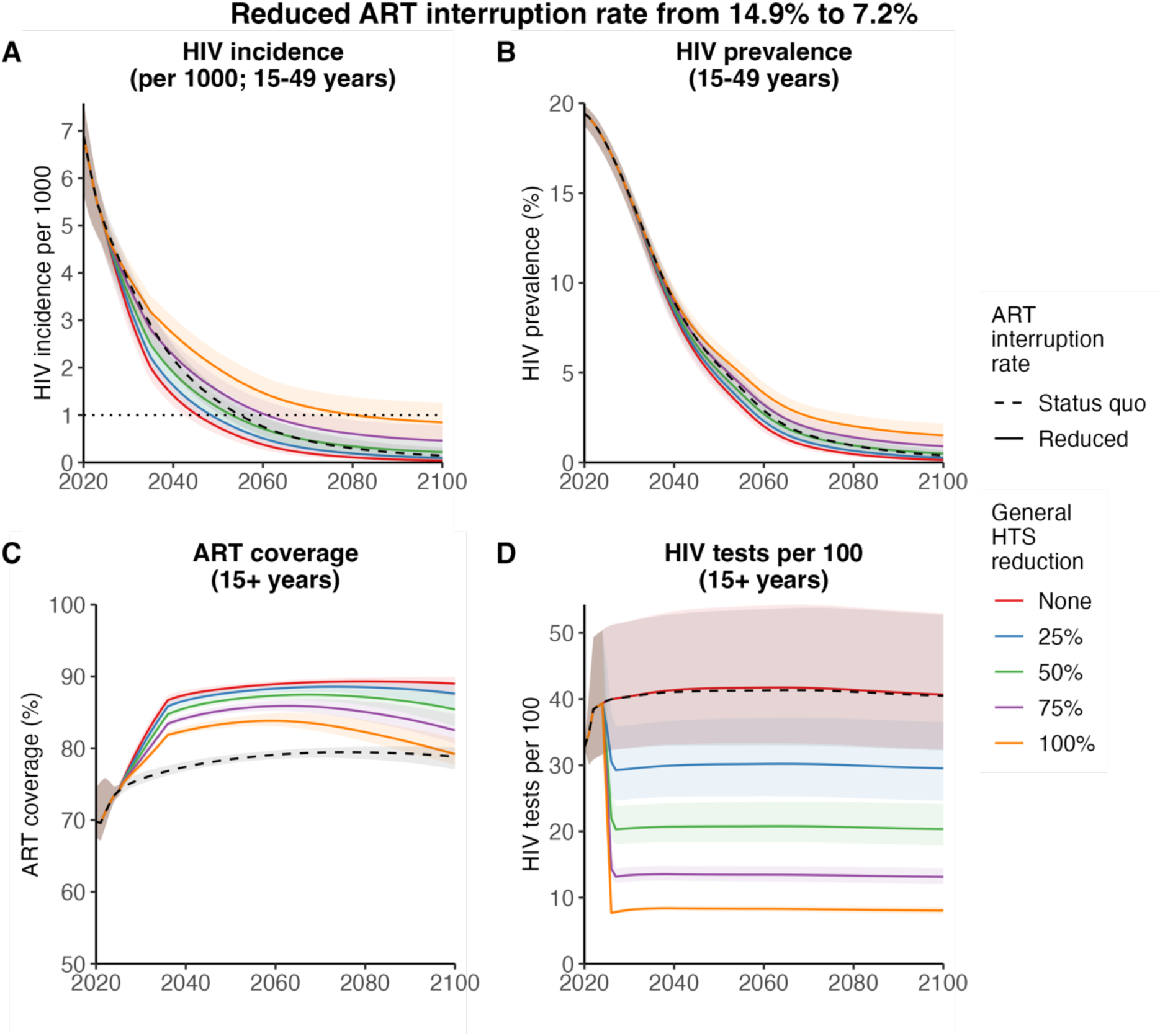
Representative scenario in which ART interruption rate reduced from 14.9% to 7.2% per year between 2025–2035. Changes in incidence, prevalence, ART coverage and HIV tests over time when general HTS was reduced, and ART interruption was reduced. Representative figures of HIV incidence rate (15–49 years) (A) per 1000 (mean and 95% CI) including an indication of when ‘virtual elimination’ (incidence <1/1000) was attained (dotted line), HIV prevalence (15–49 years) (B), ART coverage of adults (over 15 years) (C) and HIV tests per 100 (over 15 years) (D) between 2020 and 2100 showing status quo testing and treatment (ART coverage in 2035 of 76.7%), no testing reduction (ART coverage in 2035 increased to 85.8%) and general HTS reductions of 25%, 50%, 75% and 100% from 2025 (ART coverage in 2035 increased to 85.8%).

**Figure S8:**
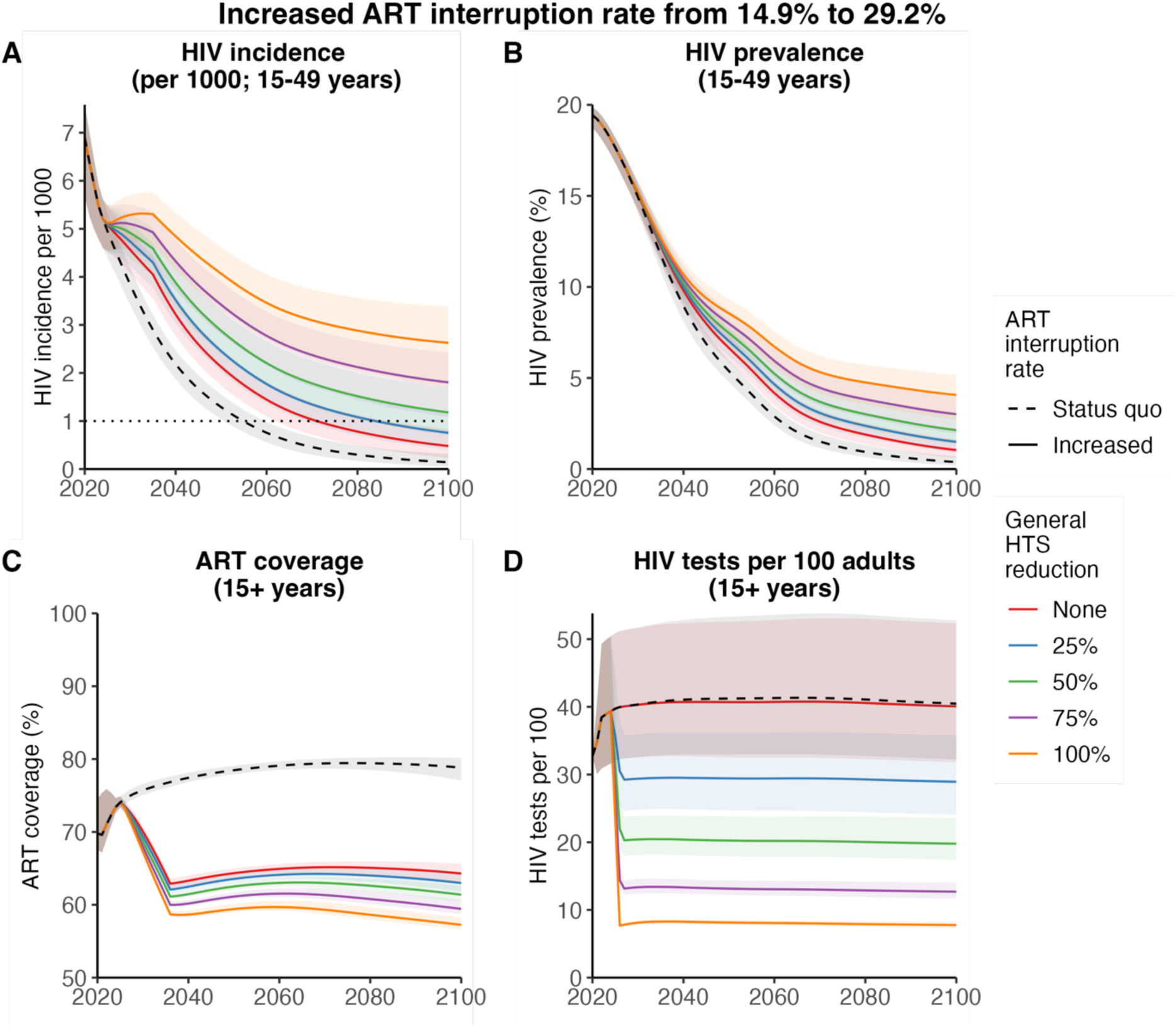
Representative scenario in which ART interruption rate increased from 14.9% to 29.2% per year between 2025–2035. Changes in incidence, prevalence, ART coverage and HIV tests over time when general HTS was reduced, and ART interruption was increased. Representative figures of HIV incidence rate (15–49 years) (A) per 1000 (mean and 95% CI) including an indication of when ‘virtual elimination’ (incidence <1/1000) was attained (dotted line), HIV prevalence (15–49 years) (B), ART coverage of adults (over 15 years) (C) and HIV tests per 100 (over 15 years) (D) between 2020 and 2100 showing status quo testing and treatment (ART coverage in 2035 of 76.7%), no testing reduction (ART coverage in 2035 decreased to 64.2%) and general HTS reductions of 25%, 50%, 75% and 100% from 2025 (ART coverage in 2035 decreased to 64.2%).

**Figure S9:**
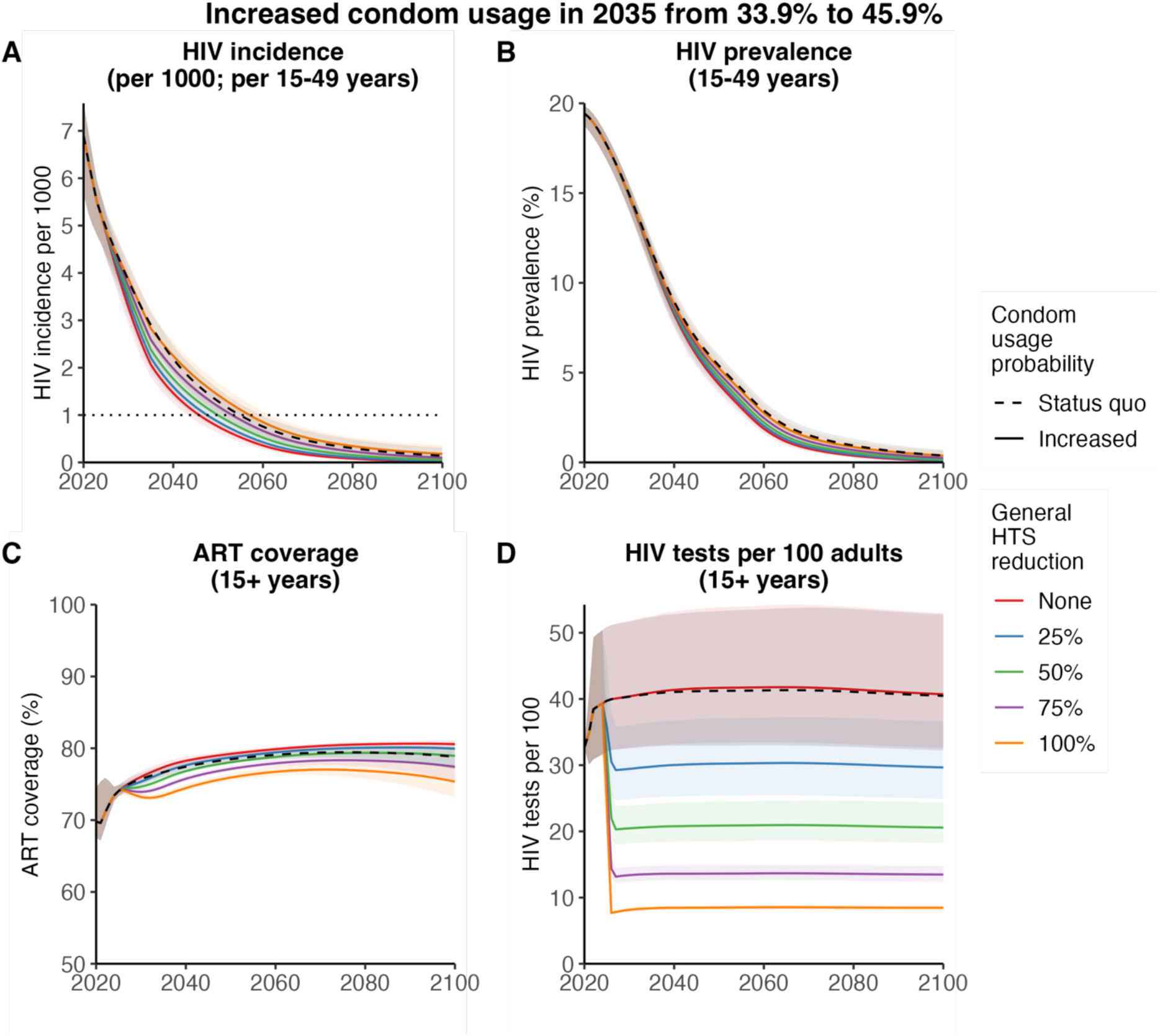
Representative scenario in which proportion of sex acts covered by condoms increased from 33.9% to 45.9% between 2025–2035. Changes in incidence, prevalence, ART coverage and HIV tests over time when general HTS was reduced, and condom usage was increased. Representative figures of HIV incidence rate (15–49 years) (A) per 1000 (mean and 95% CI) including an indication of when ‘virtual elimination’ (incidence <1/1000) was attained (dotted line), HIV prevalence (15–49 years) (B), ART coverage of adults (over 15 years) (C) and HIV tests per 100 (over 15 years) (D) between 2020 and 2100 showing status quo testing and condom usage (condom usage in 2035 of 33.9%), no testing reduction (condom usage in 2035 increased to 45.9%) and general HTS reductions of 25%, 50%, 75% and 100% from 2025 (condom usage in 2035 increased to 45.9%).

**Figure S10:**
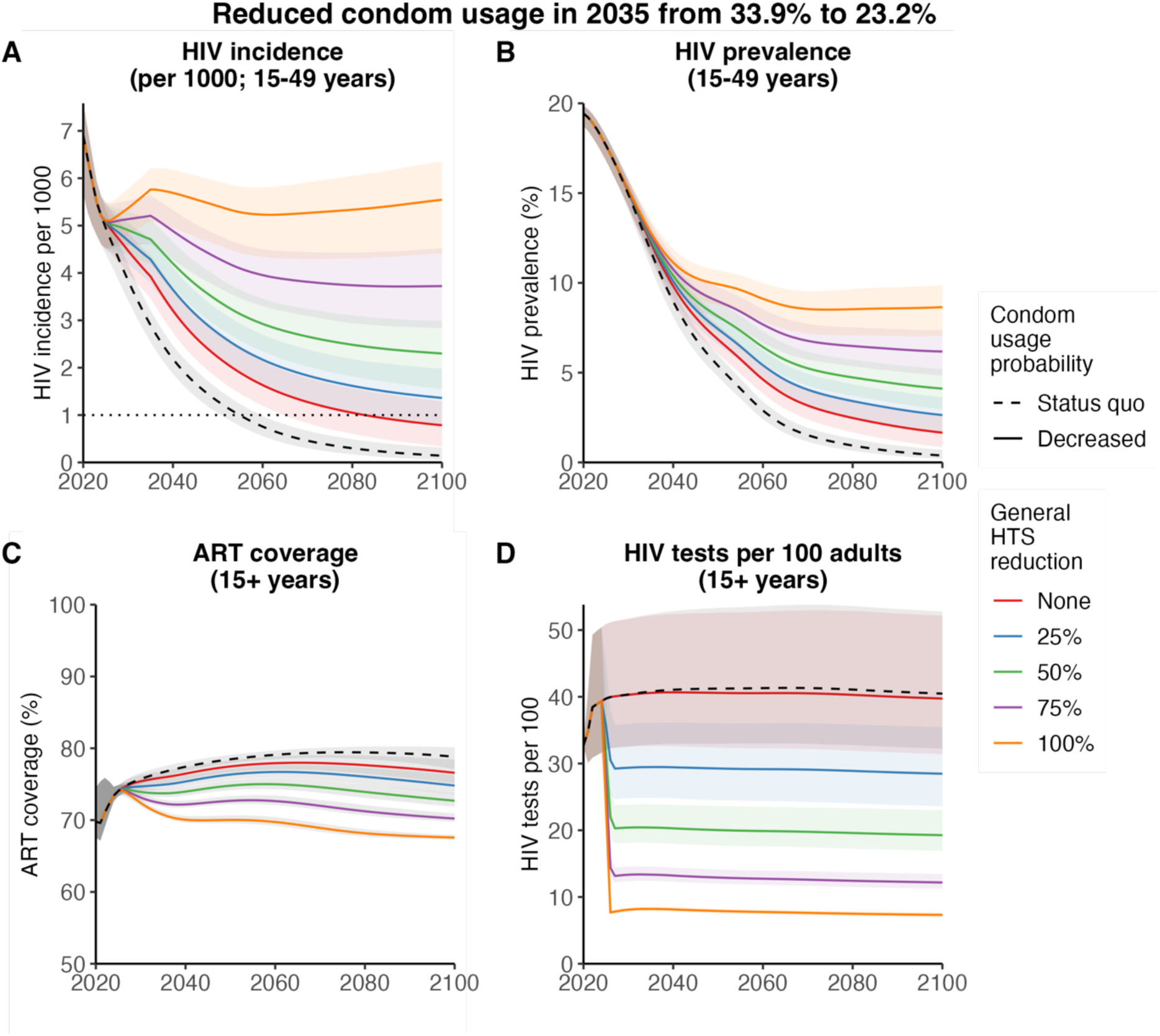
Representative scenario in which proportion of sex acts covered by condoms reduced from 33.9% to 23.2% between 2025–2035. Changes in incidence, prevalence, ART coverage and HIV tests over time when condom usage was decreased. Representative figures of HIV incidence rate (15–49 years) (A) per 1000 (mean and 95% CI) including an indication of when ‘virtual elimination’ (incidence <1/1000) was attained (dotted line), HIV prevalence (15–49 years) (B), ART coverage of adults (over 15 years) (C) and HIV tests per 100 (over 15 years) (D) between 2020 and 2100 showing status quo testing and condom usage (condom usage in 2035 of 33.9%), no testing reduction (condom usage in 2035 decreased to 23.2%) and general HTS reductions of 25%, 50%, 75% and 100% from 2025 (condom usage in 2035 decreased to 23.2%).

**Figure S11:**
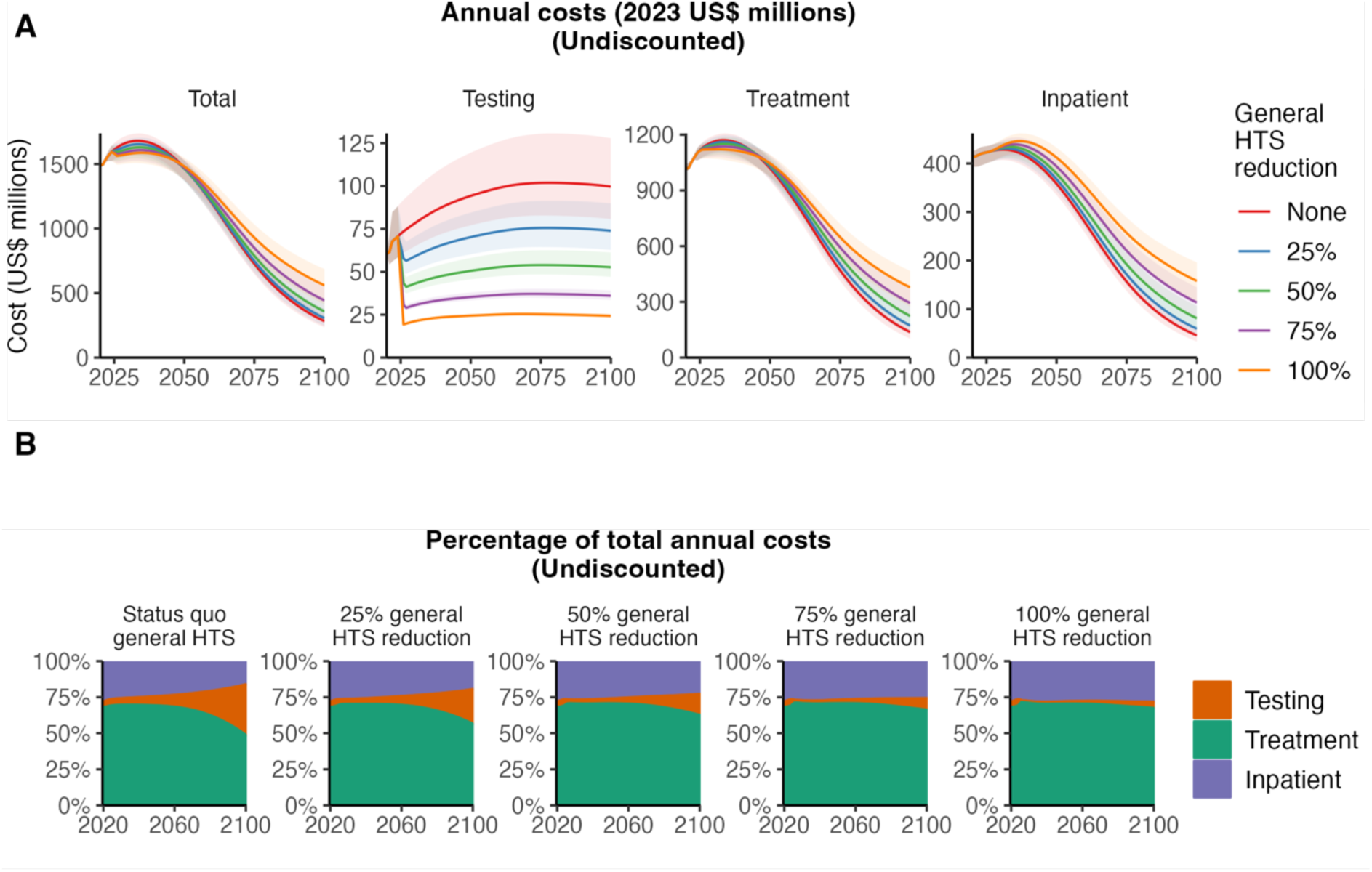
Annual HIV testing and treatment programme costs by category over 2025–2100. (A) Annual cost for total HIV testing and treatment programme and cost for testing, treatment, and care under status quo scenario and strategies reducing general HTS by 25%, 50% 75%, and 100%. (B) Distribution of total testing and treatment programme cost towards HIV testing, treatment, and care over time under each scenario.

**Figure S12.**
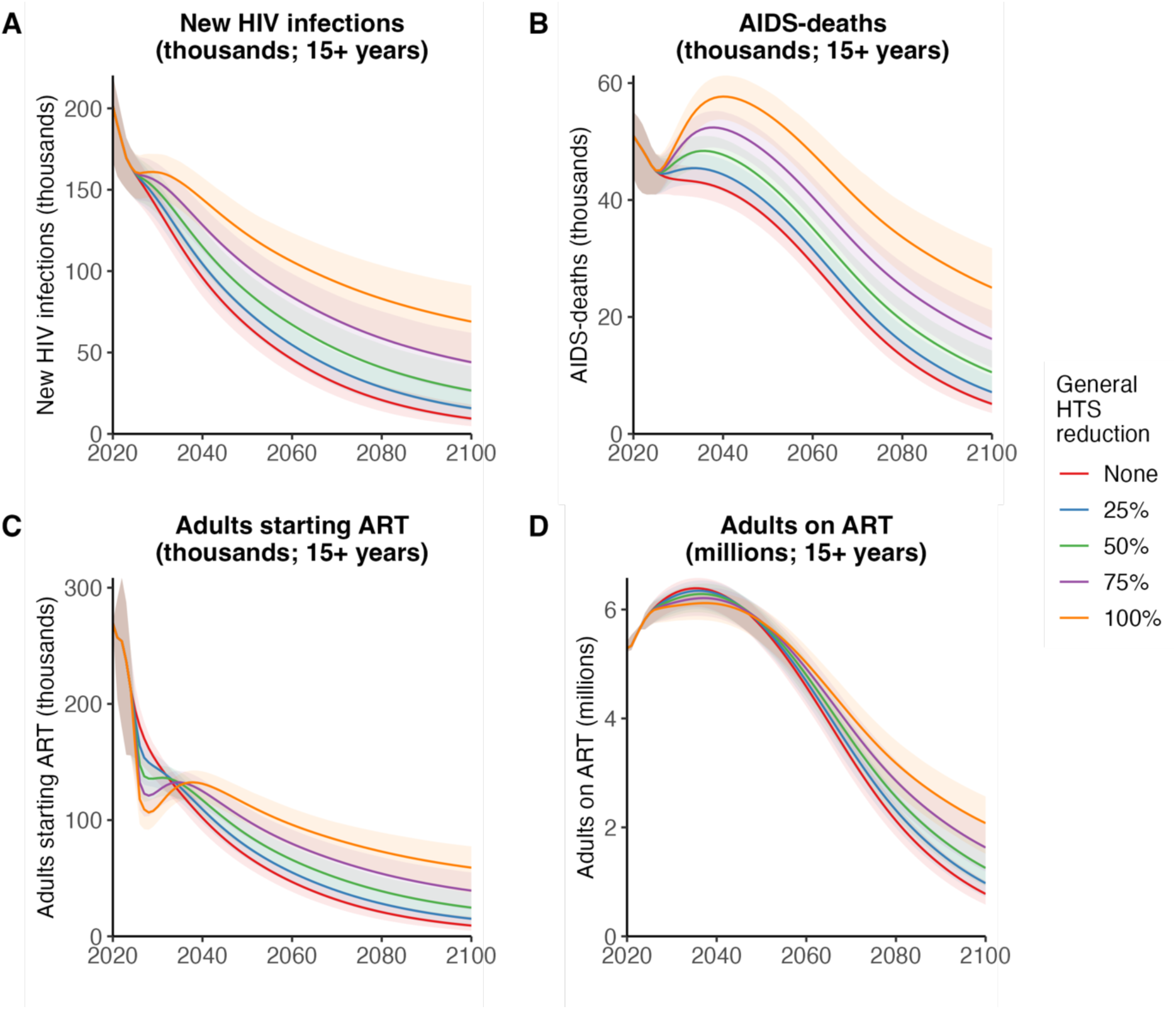
Changes in new HIV infections, AIDS-deaths, adults starting ART and adults on ART over time when general HTS was reduced. Numbers of (A) New HIV infections (over 15 years), (B) AIDS-related deaths (over 15 years), (C) adults starting ART (over 15 years), and (D) total adults on ART (over 15 years) between 2020 and 2100. Figures represent status quo scenario (no testing reduction) and general HTS reductions of 25%, 50%, 75% and 100% implemented in 2025. Lines represent posterior mean and shaded areas are 95% credible intervals.

## References

1. UNAIDS. Understanding fast-track: accelerating action to end the AIDS epidemic by 2030. Geneva, Sweden: Joint United Nations Programme on HIV/AIDS; 2015. Accessed on: 02 May 2023. Available from: https://www.unaids.org/en/resources/documents/2015/201506_JC2743_Understanding_FastTrack.

2. UNAIDS. The path that ends AIDS: UNAIDS Global AIDS Update 2023. Geneva, Sweden: Joint United Nations Programme on HIV/AIDS; 2023. Accessed on: 24 July 2023. Available from: https://www.unaids.org/en/resources/documents/2023/global-aids-update-2023.

3. PEPFAR. Fullling America’s Promise to End the HIV/AIDS Pandemic by 2030. Washington, DC: United States President’s Plan for AIDS Relief; 2022. Accessed on: 24 July 2023. Available from: https://www.state.gov/wp-content/uploads/2022/11/PEPFARs-5-Year-Strategy_WAD2022_FINAL_COMPLIANT_3.0.pdf.

4. Stover J, Bollinger L, Izazola JA, Loures L, Delay P, Ghys PD. What Is Required to End the AIDS Epidemic as a Public Health Threat by 2030? The Cost and Impact of the Fast-Track Approach. PLOS ONE. 2016;11(5):e0154893.

5. UNAIDS. AIDSinfo: Global data on HIV epidemiology and response. Joint United Nations Programme on HIV/AIDS 2023. Accessed on: 02 August 2023. Available from: https://aidsinfo.unaids.org/.

6. UNAIDS. UNAIDS data 2022. Geneva, Sweden: Joint United Nations Programme on HIV/AIDS; 2022. Accessed on: 13 February 2023. Available from: https://www.unaids.org/en/resources/documents/2023/2022_unaids_data.

7. Johnson LF, Van Rensburg C, Govathson C, Meyer-Rath G. Optimal HIV testing strategies for South Africa: a model-based evaluation of population-level impact and cost-effectiveness. Scientific Reports. 2019;9(1).

8. Johnson LF, Hallett TB, Rehle TM, Dorrington RE. The effect of changes in condom usage and antiretroviral treatment coverage on human immunodeficiency virus incidence in South Africa: a model-based analysis. Journal of The Royal Society Interface. 2012;9(72):1544–54.

9. Johnson LF, May MT, Dorrington RE, Cornell M, Boulle A, Egger M, et al. Estimating the impact of antiretroviral treatment on adult mortality trends in South Africa: A mathematical modelling study. PLoS Med. 2017;14(12):e1002468.

10. Granich R, Gupta S, Hersh B, Williams B, Montaner J, Young B, et al. Trends in AIDS Deaths, New Infections and ART Coverage in the Top 30 Countries with the Highest AIDS Mortality Burden; 1990–2013. PLOS ONE. 2015;10(7):e0131353.

11. Johnson LF, Dorrington RE. Thembisa version 4.5: A model for evaluating the impact of HIV/AIDS in South Africa. 2022. Accessed on: 17 Dec 2023. Available from: https://www.thembisa.org/.

12. Granich RM, Gilks CF, Dye C, De Cock KM, Williams BG. Universal voluntary HIV testing with immediate antiretroviral therapy as a strategy for elimination of HIV transmission: a mathematical model. The Lancet. 2009;373(9657):48-57.

13. Meyer-Rath G, Brennan AT, Fox MP, Modisenyane T, Tshabangu N, Mohapi L, et al. Rates and cost of hospitalisation before and after initiation of antiretroviral therapy in urban and rural settings in South Africa. Journal of acquired immune deficiency syndromes (1999). 2013;62(3):322.

14. PEPFAR. PEPFAR 2022 Country and Regional Operational Plan (COP/ROP) Guidance for all PEPFAR-Supported Countries. United States President’s Emergency Plan for AIDS Relief (PEPFAR); 2022. Accessed on: 13 February 2023. Available from: https://www.state.gov/2022-country-operational-plan-guidance/.

15. Chamie G, Hickey MD, Kwarisiima D, Ayieko J, Kamya MR, Havlir DV. Universal HIV Testing and Treatment (UTT) Integrated with Chronic Disease Screening and Treatment: the SEARCH study. Current HIV/AIDS Reports. 2020;17(4):315–23.

16. Bekker L-G, Alleyne G, Baral S, Cepeda J, Daskalakis D, Dowdy D, et al. Advancing global health and strengthening the HIV response in the era of the Sustainable Development Goals: the International AIDS Society—Lancet Commission. The Lancet. 2018;392(10144):312–58.

17. Grimsrud A, Wilkinson L, Ehrenkranz P, Behel S, Chidarikire T, Chisenga T, et al. The future of HIV testing in eastern and southern Africa: Broader scope, targeted services. PLOS Medicine. 2023;20(3):e1004182.

18. Ong JJ, Coulthard K, Quinn C, Tang MJ, Huynh T, Jamil MS, et al. Risk-Based Screening Tools to Optimise HIV Testing Services: a Systematic Review. Current HIV/AIDS Reports. 2022;19(2):154–65.

19. Osler M, Hilderbrand K, Goemaere E, Ford N, Smith M, Meintjes G, et al. The Continuing Burden of Advanced HIV Disease Over 10 Years of Increasing Antiretroviral Therapy Coverage in South Africa. Clinical Infectious Diseases. 2018;66(suppl_2):S118–S25.

20. Jewell BL, Smith JA, Hallett TB. Understanding the impact of interruptions to HIV services during the COVID-19 pandemic: A modelling study. EClinicalMedicine. 2020;26:100483.

21. Hoornenborg E, Coyer L, Achterbergh RCA, Matser A, Schim Van Der Loeff MF, Boyd A, et al. Sexual behaviour and incidence of HIV and sexually transmitted infections among men who have sex with men using daily and event-driven pre-exposure prophylaxis in AMPrEP: 2 year results from a demonstration study. The Lancet HIV. 2019;6(7):e447–e55.

22. Ayerdi Aguirrebengoa O, Vera García M, Arias Ramírez D, Gil García N, Puerta López T, Clavo Escribano P, et al. Low use of condom and high STI incidence among men who have sex with men in PrEP programs. PLoS One. 2021;16(2):e0245925.

23. Johnson LF, Chiu C, Myer L, Davies M-A, Dorrington RE, Bekker L-G, et al. Prospects for HIV control in South Africa: a model-based analysis. Global Health Action. 2016;9(1):30314.

24. Hontelez JAC, Lurie MN, Bärnighausen T, Bakker R, Baltussen R, Tanser F, et al. Elimination of HIV in South Africa through Expanded Access to Antiretroviral Therapy: A Model Comparison Study. PLoS Medicine. 2013;10(10):e1001534.

25. Stone J, Mukandavire C, Boily MC, Fraser H, Mishra S, Schwartz S, et al. Estimating the contribution of key populations towards HIV transmission in South Africa. Journal of the International AIDS Society. 2021;24(1).

26. Garnett GP. Reductions in HIV incidence are likely to increase the importance of key population programmes for HIV control in sub-Saharan Africa. Journal of the International AIDS Society. 2021;24(S3).

27. Jamieson L, Johnson LF, Matsimela K, Sande LA, D’Elbée M, Majam M, et al. The cost effectiveness and optimal configuration of HIV self-test distribution in South Africa: a model analysis. BMJ Global Health. 2021;6(Suppl 4):e005598.

28. Jamieson L, Johnson LF, Nichols BE, Delany-Moretlwe S, Hosseinipour MC, Russell C, et al. Relative cost-effectiveness of long-acting injectable cabotegravir versus oral pre-exposure prophylaxis in South Africa based on the HPTN 083 and HPTN 084 trials: a modelled economic evaluation and threshold analysis. The Lancet HIV. 2022;9(12):e857–e67.

29. Meyer-Rath G., Jamieson L., Johnson L. South African HIV Investment Case - Full Report. 2021. Accessed on: 18 July 2023. Available from: https://www.heroza.org/wp-content/uploads/2021/12/HIV-Investment-Case-2021-Full-report-final.pdf.

30. Akullian A, Morrison M, Garnett GP, Mnisi Z, Lukhele N, Bridenbecker D, et al. The effect of 90-90-90 on HIV-1 incidence and mortality in eSwatini: a mathematical modelling study. The Lancet HIV. 2020;7(5):e348–e58.

31. Eaton JW, Johnson LF, Salomon JA, Bärnighausen T, Bendavid E, Bershteyn A, et al. HIV Treatment as Prevention: Systematic Comparison of Mathematical Models of the Potential Impact of Antiretroviral Therapy on HIV Incidence in South Africa. PLoS Medicine. 2012;9(7):e1001245.

32. Jooste S, Mabaso M, Taylor M, North A, Tadokera R, Simbayi L. Trends and determinants of ever having tested for HIV among youth and adults in South Africa from 2005–2017: Results from four repeated cross-sectional nationally representative household-based HIV prevalence, incidence, and behaviour surveys. PLOS ONE. 2020;15(5):e0232883.

33. Kaplan SR, Oosthuizen C, Stinson K, Little F, Euvrard J, Schomaker M, et al. Contemporary disengagement from antiretroviral therapy in Khayelitsha, South Africa: A cohort study. PLoS Med. 2017;14(11):e1002407.

34. Johnson LF, Meyer-Rath G, Dorrington RE, Puren A, Seathlodi T, Zuma K, et al. The effect of HIV programs in South Africa on national HIV incidence trends, 2000–2019. JAIDS Journal of Acquired Immune Deficiency Syndromes. 2022;90(2):115–23.

35. Connolly C, Simbayi LC, Shanmugam R, Nqeketo A. Male circumcision and its relationship to HIV infection in South Africa: results of a national survey in 2002. South African Medical Journal. 2008;98(10):789–94.

36. Hanscom B, Janes HE, Guarino PD, Huang Y, Brown ER, Chen YQ, et al. Brief Report: Preventing HIV-1 Infection in Women Using Oral Preexposure Prophylaxis: A Meta-analysis of Current Evidence. JAIDS Journal of Acquired Immune Deficiency Syndromes. 2016;73(5):606–8.

37. Molina J-M, Capitant C, Spire B, Pialoux G, Cotte L, Charreau I, et al. On-Demand Preexposure Prophylaxis in Men at High Risk for HIV-1 Infection. New England Journal of Medicine. 2015;373(23):2237–46.

38. McCormack S, Dunn DT, Desai M, Dolling DI, Gafos M, Gilson R, et al. Pre-exposure prophylaxis to prevent the acquisition of HIV-1 infection (PROUD): effectiveness results from the pilot phase of a pragmatic open-label randomised trial. The Lancet. 2016;387(10013):53–60.

39. Meyer-Rath G, Van Rensburg C, Chiu C, Leuner R, Jamieson L, Cohen S. The per-patient costs of HIV services in South Africa: Systematic review and application in the South African HIV Investment Case. PLOS ONE. 2019;14(2):e0210497.

